# Identification of transmission clusters with high influence on shared parameter estimates in Bayesian phylodynamics

**DOI:** 10.1101/2025.11.18.25340546

**Authors:** Ariane Weber, Ruth Boersma, Sebastian Duchene

## Abstract

Estimating a shared rate of transmission from multiple clusters of pathogen genome sequences has become increasingly common. Such inferences are appealing, as they allow the overall characterisation of a polyphyletic population and the inclusion of clusters that could not easily be analysed by themselves, for example due to a very small number of sequence samples. Differences in sample size, however, can result in substantial variations in the information content each cluster carries about the rate of transmission. When the clusters additionally differ in their respective transmission rates, the shared estimate would not represent all clusters equally well. It is therefore integral to assess the influence of each cluster on the estimate. Here, we focus on Bayesian phylodynamic inference which combines an evolutionary with an epidemiological model to infer transmission rates from clusters of genetic sequences. We build on results about case influence in Bayesian models to suggest a computationally inexpensive measure that quantifies influence in the context of inference from multiple conditionally independent clusters, regardless of the exact epidemiological model. We further analyse clusters simulated under various birth-death-sampling models to evaluate the performance of the method and explore which properties can generally be drivers of high influence. We find the influence to be a complex interplay of size, persistence and rate relative to all included clusters. We further highlight that the shared estimate from clusters of different rates does not correspond to a population mean or median, but rather to a certainty-weighted average. Finally, we demonstrate the practical insights that can be gained through influence assessment in the analysis of SARS-CoV-2 genomes, sampled in Germany before the implementation of the first nation-wide lockdown in early 2020. Our results illustrate how variable the influence of individual clusters can be and how quantifying it can guide further studies, for example into transmission rate differences.

## 1 Introduction

Bayesian inference refers to the estimation of parameters-of-interest in terms of probability distributions from observed data. Given the data, these distributions directly quantify the uncertainty inherent in the estimation. To construct them, a probability model that connects the observed data with the parameters-of-interest and prior information on them is needed. As hierarchical dependencies can be incorporated into this model, Bayesian inference is a flexible method that allows complex model structures (Gelman et al., 2013). In phylogenetics, it has thus proven to be a valuable addition to other inference methods, like maximum likelihood and maximum parsimony (Felsenstein, 2004; Yang & Rannala, 2012). Bayesian phylogenetics aims to reconstruct phylogenetic trees and evolutionary parameters from genetic sequences. It has provided insights into many diversification histories on different time scales, from reconstructed evolutionary relationships of mammals (Álvarez-Carretero et al., 2021) reaching back millions of years or of human pathogens like HBV (Kocher et al., 2021) over the last ten thousand years, to the emergence of human adapted SARS-CoV-2 on the scale of years and months (Pekar, Lytras, et al., 2025).

Bayesian phylodynamics makes use of the hierarchical model structures that can be incorporated into the analyses and constructs the prior probability distribution for the phylogenetic tree from a population dynamic process. Through this approach it does not only provide a mechanistic description of the generative process of the phylogenetic tree, but in turn also connects it with an underlying population (E. M. Volz et al., 2013). Thus, in addition to the tree, population dynamic parameters can be inferred from models based on coalescent theory (Kingman, 1982) or birth-death processes (Kendall, 1948). These parameters include population sizes through time or epidemiological quantities, for example the growth rate or the reproductive number, *R*, which represents the average number of infections caused by one case. In recent years, and especially during the COVID-19 pandemic, the construction and application of models for disease transmission has grown into a focal point of Bayesian phylodynamic inference. These models can serve as powerful additions to other data-driven methods, like incidence time series analyses. Flexible coalescent models that can be adjusted to the specific epidemiological contexts are increasingly used and have, for example, revealed transmission rate heterogeneity between clusters of SARS-CoV-2 (E. Volz et al., 2021; E. M. Volz & Siveroni, 2018; Wilinski et al., 2024). Similarly, birth-death models provide an adjustable framework such that they can either be combined with or used to infer prevalence and incidence (Kühnert et al., 2014; T. G. Vaughan & Stadler, 2025; Zarebski et al., 2025).

However, infectious disease outbreaks often cannot be connected by a single phylogenetic tree to estimate the parameters-of-interest: multiple jumps of a pathogen into the host population, multiple introductions of it into the area of interest or the re-emergence of a genetic variant that impacts transmissibility result in outbreaks with complex substructure. Although substructure-aware models exist, their substantially higher computational demand restricts their practical use. Simpler models have therefore been applied more extensively. If, for example, the subpopulations form monophyletic groups, they can be joined in one analysis by assuming them to be independent realisations of the same process. Thus multiple trees that share the same parameters but are assumed to evolve independently are estimated, one for each subpopulation. This framework has been applied extensively to characterise the dispersal of several pathogens in different epidemic settings: the joint analysis of epidemiological clusters of HIV-1 infections sampled in Botswana enabled the estimation of individual reproductive numbers for each cluster by sharing the duration of infectiousness and the evolutionary rate (Novitsky et al., 2015). Inferences from genetic sequences of influenza virus in Basel, Switzerland, pointed to a correlation between infectivity and temperature (Müller et al., 2020). Through the inference of a shared growth rate from mpox virus clusters in New York City, the epidemic has been shown to follow similar transmission patterns as other sexually transmitted diseases (Pekar, Wang, et al., 2025). By combining multiple independent introductions of SARS-CoV-2 into Switzerland in 2020 the impact of non-pharmaceutical interventions on its spread has been evaluated (Nadeau et al., 2023). Several other studies have focused on the transmission of SARS-CoV-2 or HIV in specific regions, for example Washington State, Russia, Portugal and Germany (Kühnert et al., 2018; Müller et al., 2021; Safina et al., 2022; M. R. Smith et al., 2021; Vasylyeva et al., 2019; Weber et al., 2023), as well as other pathogens (Walas et al., 2024). Besides facilitating the inference of a shared parameter that is informed by multiple genetic clusters, this approach also comes with the advantage that subpopulations of smaller sizes can be included in the analysis. It has therefore not only been applied in a Bayesian setting, but also in maximum likelihood inference (Zhukova et al., 2023).

There are two main limitations for the multi-cluster framework described above: first, the individual subpopulations have to be known prior to the Bayesian inference. As this information often does not exist independently, the data has to be preprocessed to identify the subpopulations, for example through phylogeographic approaches or genetic cluster thresholds (Nadeau et al., 2023; Novitsky et al., 2015; Weber et al., 2023). Second, the combined clusters differ, often substantially, in the characteristics that inform their tree-generating distribution and therefore also the inferred parameter. Such characteristics include the number of genetic sequences or the persistence time of the subpopulation. To assess the robustness of the shared estimate, analyses on reduced data sets or with different prior assumptions have been used (Översti et al., 2025). However, repeating the inference multiple times comes with high computational costs, particularly for data sets comprising many hundreds to thousands of genome sequences. In addition, a small number of sensitivity analyses does not provide a more general understanding of the way in which specific characteristics influence the shared estimate. This becomes particularly relevant when clusters that are characterised by different rates are combined into one analysis: without a comprehensive assessment of the influence of each cluster, the shared parameter estimate cannot be considered representative of all individual clusters.

From a theoretical perspective, case influence in Bayesian inference has been studied widely (Bradlow & Zaslavsky, 1997; Cook, 1986; Danilevicz & Ehlers, 2020; McCulloch, 1989; Millar & Stewart, 2007; Peruggia, 1997; Thomas et al., 2018). Several procedures have been established based on different approaches and formal definitions of influence. Fundamentally, it is also closely related to the problem of quantifying predictive accuracy of Bayesian models (see chapter 6 in McElreath, 2020 and Vehtari et al., 2016). Leave-one-out strategies or case deletion analyses, for example, study the behaviour of the shared estimate after removal of individual or groups of cases from the observed data set (Peruggia, 1997; Weiss & Cook, 1992). Influence is then understood as the change induced in the posterior of the shared estimate when the information provided by that case is completely neglected. Practical implementations can differ in how this change is quantified and in how the case-deleted posterior distributions are calculated. Besides analytical solutions, the posterior distributions can be determined via independent approximations through Markov chain Monte Carlo (MCMC) sampling or by reweighting of the posterior sample of the full data set. While running MCMC chains is computationally intensive, reweighting approaches like importance sampling can lack in accuracy, particularly when the influence of a case is high (Peruggia, 1997). To quantify difference in the sampled posterior distributions, point estimates like the first moment are, for example, compared in their absolute value or distances between the full posterior distributions are calculated, e.g. the Wasserstein distance. Alternatively to case-deletion approaches, the impact of slight perturbations in the likelihood function of the respective case can be considered: local influence techniques evaluate the distance between the full posterior distribution and the weighted version of it in the area around equality (McCulloch, 1989). To quantify the distance, basic divergence measures like the Kullback-Leibler or Bregman divergence have been used, but others can equally be applied (Danilevicz & Ehlers, 2020; Millar & Stewart, 2007).

To assess the local influence of genetic clusters on a shared parameter in a Bayesian phylo-dynamic analysis, we focus in this study on the approach based on a geometric weighting of the likelihood function, introduced by Millar and Stewart, 2007 and revisited by Thomas et al., 2018. We will first summarise the theoretical background and set out how it extends to Bayesian phylo-dynamic analysis. From this, we will suggest a simple measure that can be used to evaluate the influence each cluster has on the joint analysis, independent of the exact form of the probability distributions for the data and phylogenetic tree. We will then turn our focus to birth-death-sampling models as tree-generating processes and the inference of one shared transmission rate jointly from multiple genetic clusters using the software BEAST2 (Bouckaert et al., 2014, 2019). Through the analysis of simulated data sets, we explore how cluster size, persistence and rate can practically determine the influence of a cluster. To demonstrate their complex interplay we end with presenting the analysis of SARS-CoV-2 genome sequences, which were collected in Germany before the implementation of the first nation-wide large-scale non-pharmaceutical intervention during the COVID-19 pandemic.

## 2 Results

### 2.1 Local influence in Bayesian phylodynamics

#### Theoretical framework

We address the problem of inferring a shared parameter ̂θ from a collection of *n* multiple sequence alignments (MSAs) 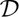 = {*D_i_* | *i* = 1*, . . ., n*}. Each alignment *D_i_* represents a genetic cluster composed of a set of genetic sequences. The sequences in *D_i_* correspond to the tips of a bifurcating tree *T_i_* ∈ 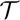 = {*T_i_* | *i* = 1*, . . ., n*}. In a Bayesian inference framework we are interested in the posterior distribution:

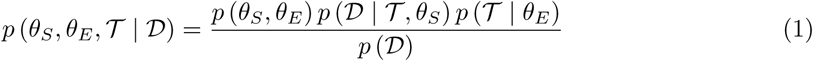

where 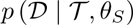 is the likelihood of 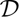 defined by an evolutionary model on 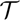, parameterised by *θ_S_*, and 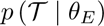 is the probability of the trees calculated from a population dynamic model with parameters *θ_E_*. Assuming conditional independence of the genetic clusters in their evolutionary as well as population process, meaning that the elements of both 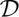 and 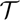 are independent identically distributed (i.i.d.), their likelihoods and tree prior distributions can be written as products over all clusters, i.e.

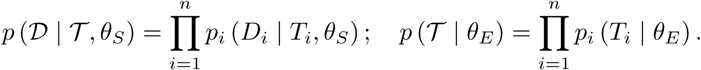

In the following sections, we assume *p_i_* (*D_i_* | *T_i_, θ_S_*) to be the phylogenetic likelihood of tree *T_i_*, as described by Felsenstein (Felsenstein, 1981), and *p_i_* (*T_i_* | *θ_E_*) to be derived from a birth-death-sampling model generating the phylogenetic tree *T_i_*, formulated by Stadler et al., 2012. However, other formulations, like more complex birth-death models (MacPherson et al., 2021) or coalescent-based probability distributions for the tree (Kingman, 1982), can be used analogically. Depending on the specific research question, the parameter of interest ̂θ can correspond to any parameter that is shared across the processes underlying the observed data. Here, we focus on the transmission rate due to its epidemiological relevance and high prevalence in the existing relevant literature (Müller et al., 2021; Nadeau et al., 2023; Vasylyeva et al., 2019; Weber et al., 2023). Thus, we assume ̂θ to be the transmission rate (also: lambda), which is one of the parameters of the population dynamic model, and denote all remaining model parameters by *θ* for brevity. We refer to the contribution 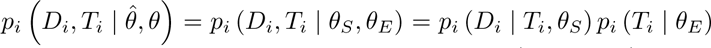 of cluster *i* to the full posterior distribution as *cluster likelihood*, such that 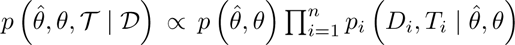 To quantify the influence of each cluster on the joint estimation of θ^, we study the extent to which small changes in its information content affect the posterior distribution. We vary the information content by applying a geometric weighting to the cluster likelihoods, thus defining the *quasi-posterior distribution*

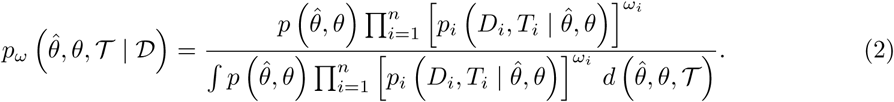

with *ω* = (*ω*_1_*, …, ω_n_*)*^T^* . Note that the integral is finite in an open neighbourhood of *ω* = *ω_eq_* = (1*, . . .,* 1)*^T^* (Appendix B in Thomas et al., 2018). We then quantify the effect of the weighting using the Kullback-Leibler divergence between the original and quasi-posterior:

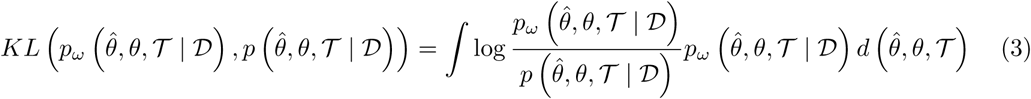

In this approach we follow Millar and Stewart, 2007, who suggested it for studying the influence of individual observations, and by Thomas et al., 2018, who extended it to all observations simultane-ously. We will therefore only shortly summarise their further argument. To quantify the influence each cluster has on the joint posterior, we consider the behaviour of *KL* (*p_ω_* (· | ·) *, p* (· | ·)) in a small neighbourhood around *ω_eq_*, i.e. the *local influence*. Through a local approximation of the Kullback-Leibler divergence in terms of a second-order Taylor series expansion and evaluation of the behaviour of the resulting Hessian matrix around *ω_eq_*, we find the curvature of the Kullback-Leibler divergence to be well approximated by the variance-covariance matrix of the logarithmic cluster likelihoods:

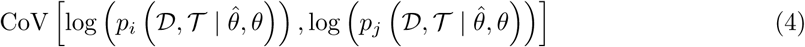

with *i, j* ∈ 1*, . . ., n*. The variance-covariance matrix of the cluster likelihoods can easily be calcu-lated using the sampled values in an MCMC analysis of the full data set. The variances, i.e. the diagonal elements of the matrix, quantify the local influence of each individual cluster on the joint analysis, while the off-diagonal elements, the covariances, reveal synergistic and directional effects, for example if two clusters pull the estimate in the same direction. In Supplementary Figure 1 and 2 we show how the variance of the cluster likelihoods, quantifying the local influence, relates to em-pirical differences between posterior distributions of cluster deletion analyses. In this comparison, we contrast both a different measure of influence (local perturbation vs. complete deletion) and a different measure of divergence between probability distributions (Kullback-Leibler divergence vs. Wasserstein distance). As such we do not necessarily expect these to yield the same values or trends; we rather illustrate how and in which cases they differ: When considering inferences from i.i.d. clusters, we find little agreement regarding the influence on the full posterior, whereas the agreement increases when considering the influence on the transmission rate only. However, we find the relationship between local influence and case-deletion to be well approximated by a linear function for analyses of non-identically distributed clusters, both for the full posterior as well as the posterior sample of the transmission rate only. In this case, the scaled variances are also substantially higher than for the i.i.d. clusters.

**Figure 1:**
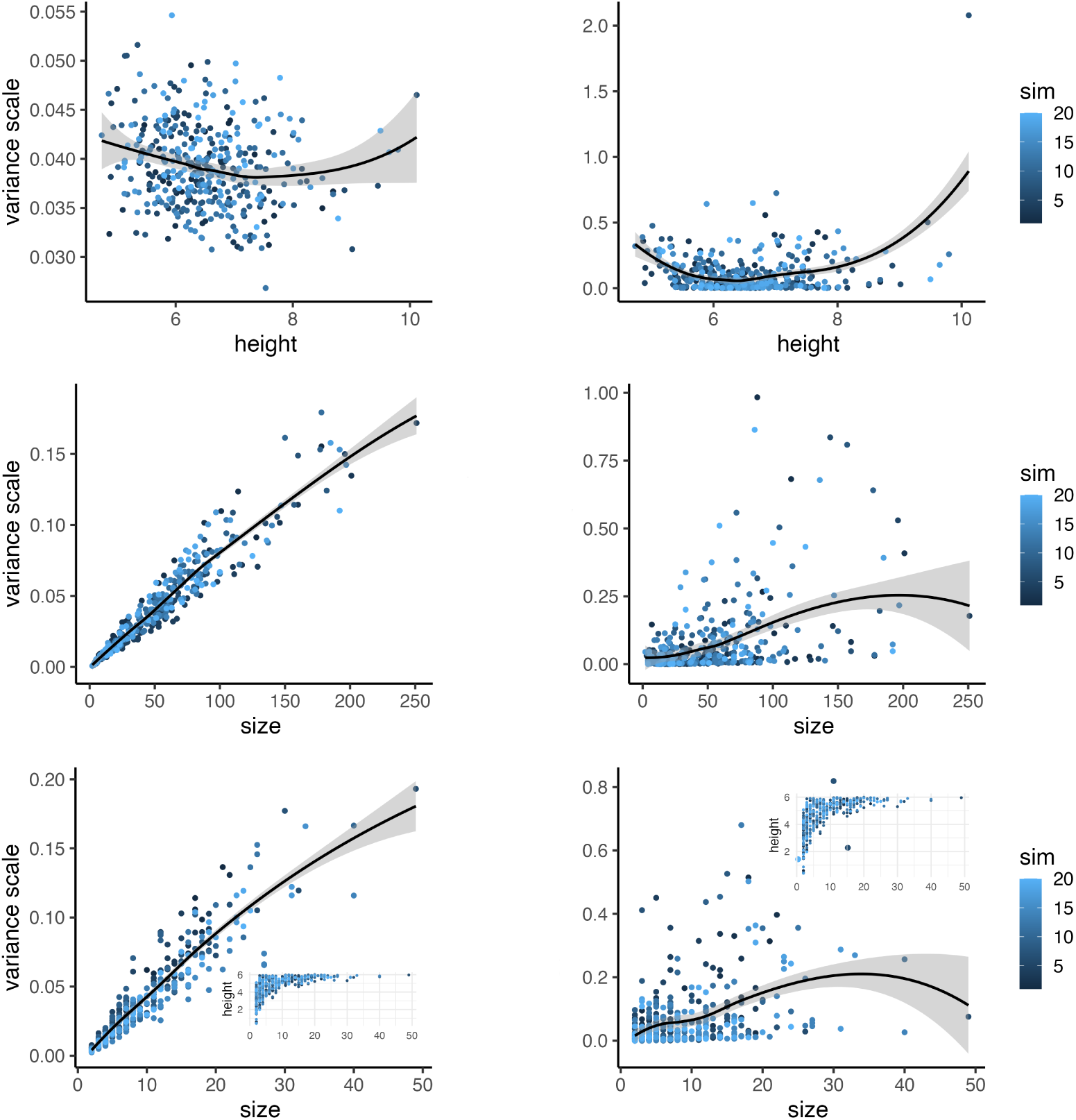
Drivers of cluster influence given independent identically distributed clusters: Scaled variance of the 20 logarithmic cluster likelihoods for each of the 20 simulation replicates against the respective cluster height or size. Simulation replicates are indicated by colour (legend ’sim’); the black line within the grey area illustrates the smoothed trend of the data and the associated error. Plots in the left panels correspond to analyses of the simulated data when tree topology, transmission rate and evolutionary rate are estimated, plots in the right panel to those estimating only the transmission rate. In the top row only trees with 20 tips are simulated, in the second row trees with a height of 3.0*y* and in the third row trees with processes running for up to 6.0*y*.

**Figure 2:**
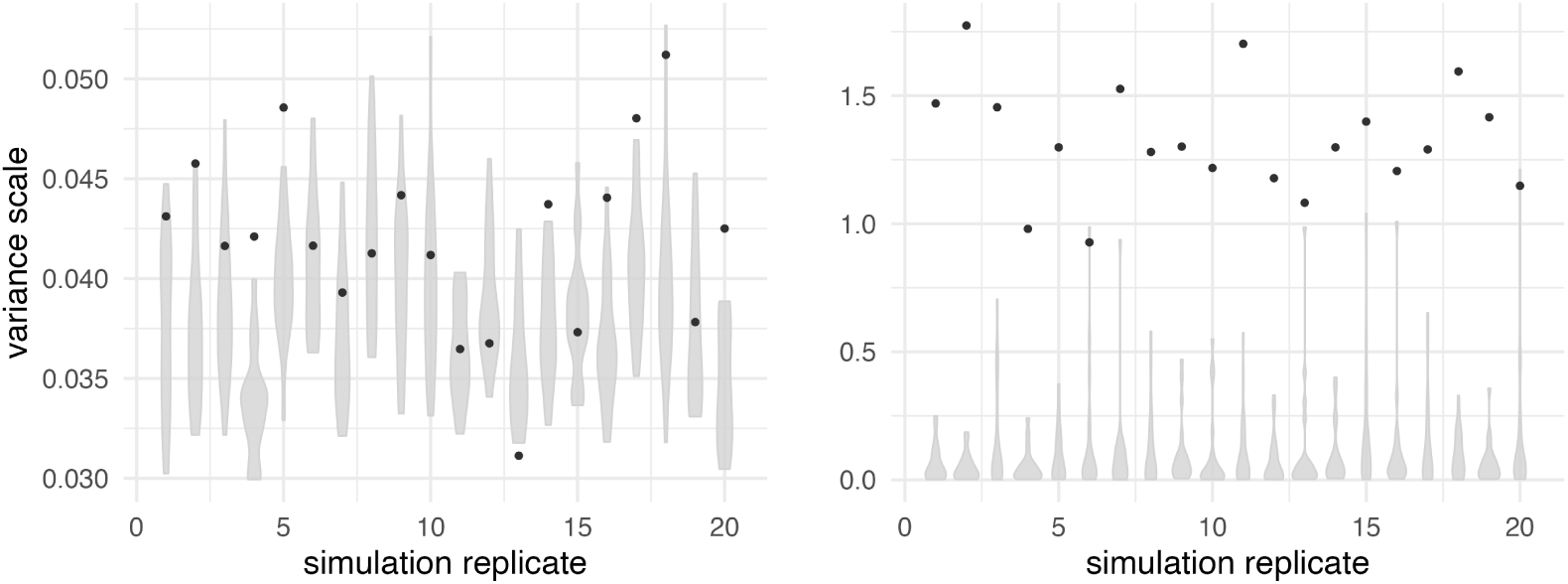
Drivers of cluster influence given a single non-identically distributed cluster: Scaled variance of the 20 logarithmic cluster likelihoods against the simulation replicates. The 19 i.i.d. clusters are represented by the violin plots, the cluster simulated under a higher transmission rate by the black dot. Plots in the left panel correspond to analyses of the simulated data when tree topology, transmission rate and evolutionary rate are estimated, plots in the right panel to estimating only the transmission rate.

#### Scaling according to model complexity

To make the simulation replicates in Section 2.2.1 comparable between each other, we follow Millar and Stewart, 2007 and work with the scaled variance

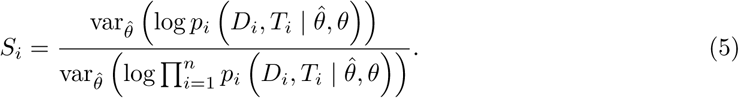

As the magnitude of the variance depends on the model complexity, using the scaled variance also allows comparison between models of different complexity, for example when a different number of parameters is estimated.

#### Combined influence of multiple clusters

Often we are not only interested in the influence each cluster has by itself, but also in the joined influence of multiple clusters. In this case the combined influence can be summarised by the variances of the respective summed up logarithmic cluster likelihoods.

#### Inference of multiple parameters

Similarly, instead of being interested in the influence of one cluster on the entire analysis, we might mainly be concerned with its influence on one specific parameter. As assessing the influence of only one of multiple estimated parameters would require integration over all other parameters, Thomas et al., 2018 propose to reevaluate the cluster like-lihoods for the posterior sample of the parameter-of-interest with all other previously estimated parameters fixed to their estimation mean or median. In Supplementary Figure 3 we provide a comparison of the approximation to the inference of only the respective parameter. As the inference of the other parameters also impacts the influence on the parameter-of-interest, we do not expect these two to be equal, but only to show similar trends, which we observe to be the case. In addition, we relate the single-parameter influence to the empirical difference induced in the posterior distribution of the single parameter under a case-deletion setting in Supplementary Figures 1 and 2. These, too, suggest that the approximation works well in the phylodynamic setting.

### 2.2 Drivers of cluster influence

Utilising the above approach, we now turn to the phylodynamic inference framework implemented in the software package BEAST2 (Bouckaert et al., 2014) to study local influence in a joint analysis of multiple genetic clusters. As we are interested in the inference of epidemiological parameters, particularly the transmission rate, we focus on the joint analysis of transmission clusters using birth-death-sampling models. This corresponds to the basic inference problem 1 with the probability of the sampled tree being calculated from the birth-death-sampling density introduced by Stadler et al., 2012. We first run basic phylodynamic inferences of the transmission rate based on MCMC sampling from the posterior distribution, to then calculate the covariance matrix of all sampled logarithmic cluster likelihoods. For the influence of single clusters, we focus on the diagonal elements, i.e. the variances, while turning to the sign of the covariance matrix to understand in which direction the respective clusters pull the estimate relative to each other.

In the above implementation, three main factors can influence the variance of the birth-death-sampling probability density: the number of branches, their lengths and the three rate parameters, i.e. those that describe birth/transmission, death and sampling intensity, individually. To study how these act on the variance individually and combined, we empirically explore the local influence as a function of tree height, as a proxy for the branch lengths, tree size, i.e. the number of samples or tips, which is a linear function of the number of internal nodes, and the transmission rate.

#### 2.2.1 Simulations under identical distribution

We consider 20 simulation replicates, in each we simulate 20 bifurcating trees from a birth-death-sampling process, controlling for either tree height, size or neither, and use nucleotide sequence alignments simulated along these trees to infer either the tree, evolutionary and transmission rate or only the transmission rate, respectively (estimates of the transmission rate are given in Supplementary table 1). In these settings, all 20 trees combined in one analysis are independent realisations of the same birth-death-sampling process, as assumed in the analytical formulation of the inference problem.

##### Impact of tree height (setting I)

In the first row of Figure 1 we show the scaled variance of the logarithmic cluster likelihoods (equation 5) plotted against the respective tree heights. Both for the inference of tree and transmission rate (left column) and of only the transmission rate (right column), we do not detect a strong association between the two quantities.

##### Impact of tree size (setting II)

The association between tree size and scaled variance is illustrated in the second row Figure 1. When estimating both tree and transmission rate (left column), the influence depends approximately linearly on the size of the cluster. This, however, is not the case when only the transmission rate is estimated: although smaller clusters tend to have less influence, bigger clusters do not necessarily have more.

##### Combined impact of tree height and size (setting III)

When both the tree size and the height are allowed to vary between clusters, we find a similar pattern. While inferences of both tree and transmission rate show a linear relationship between influence and size, we only observe a tendency for larger trees to have higher influence when inferring the transmission rate alone. As bigger trees are generally also higher (insets), we also see a tendency for higher influence of higher trees. Comparing the influence on the transmission rate between the individual effect of the tree size and the combined effect with height (second and third row, right column) suggests that, in these simulation settings, size-dependent effects on the transmission rate only become strong above sizes of 50 samples.

In summary, we find that clusters with more samples are associated with more peaked cluster likelihoods in log space and thus higher variances. However, this effect is only strongly present when the tree itself is part of the inference, indicating that the uncertainty of the cluster likelihood is to a large part mediated by uncertainty in the tree. Equally, when we consider the influence on only the rate in an analysis in which it is estimated together with the tree, larger trees tend to be associated with higher influence, although the relationship is not definitive (Supplementary Figure 3). The tree height, however, we do not find to be strongly associated with higher influence. We also note that the scaled variances observed on identically distributed trees are generally relatively small.

#### 2.2.2 Simulations under non-identical distribution

As the analysis of real-world data often combines clusters that are, other than analytically assumed, not realisations of the same birth-death-sampling process (Weber et al., 2023), we combine trees that were simulated under different transmission rates into one analysis to better understand practical applications (estimates of the transmission rate are listed in Supplementary tables 2 and 3).

##### Impact of rate outlier (setting IV)

We first consider a scenario in which one of 20 clusters grows with a higher rate. Figure 2 presents the resulting influence metric of all simulated clusters, grouped by transmission rate, against the respective simulation replicate. For the inference of both tree and transmission rate, the different transmission rate does not have a distinct effect on the variance of the posterior distribution in log space. However, on the transmission rate itself we find the cluster with higher rate to have a higher influence than the other 19 i.i.d. clusters. (see also Supplementary Figure 3).

##### Combined impact of rate and height (setting V)

Second, we evaluate how the combination of differences in rate and tree height drive the influence of individual clusters. The results are shown in the first two rows of Figure 3. When we combine all 20 clusters with rates varying between 1.6*y^−^*^1^ and 4.4*y^−^*^1^ in one inference, the posterior distribution of the rate is not centred around the mean or median of the rates that were used for simulation, rather pulled downwards.

**Figure 3:**
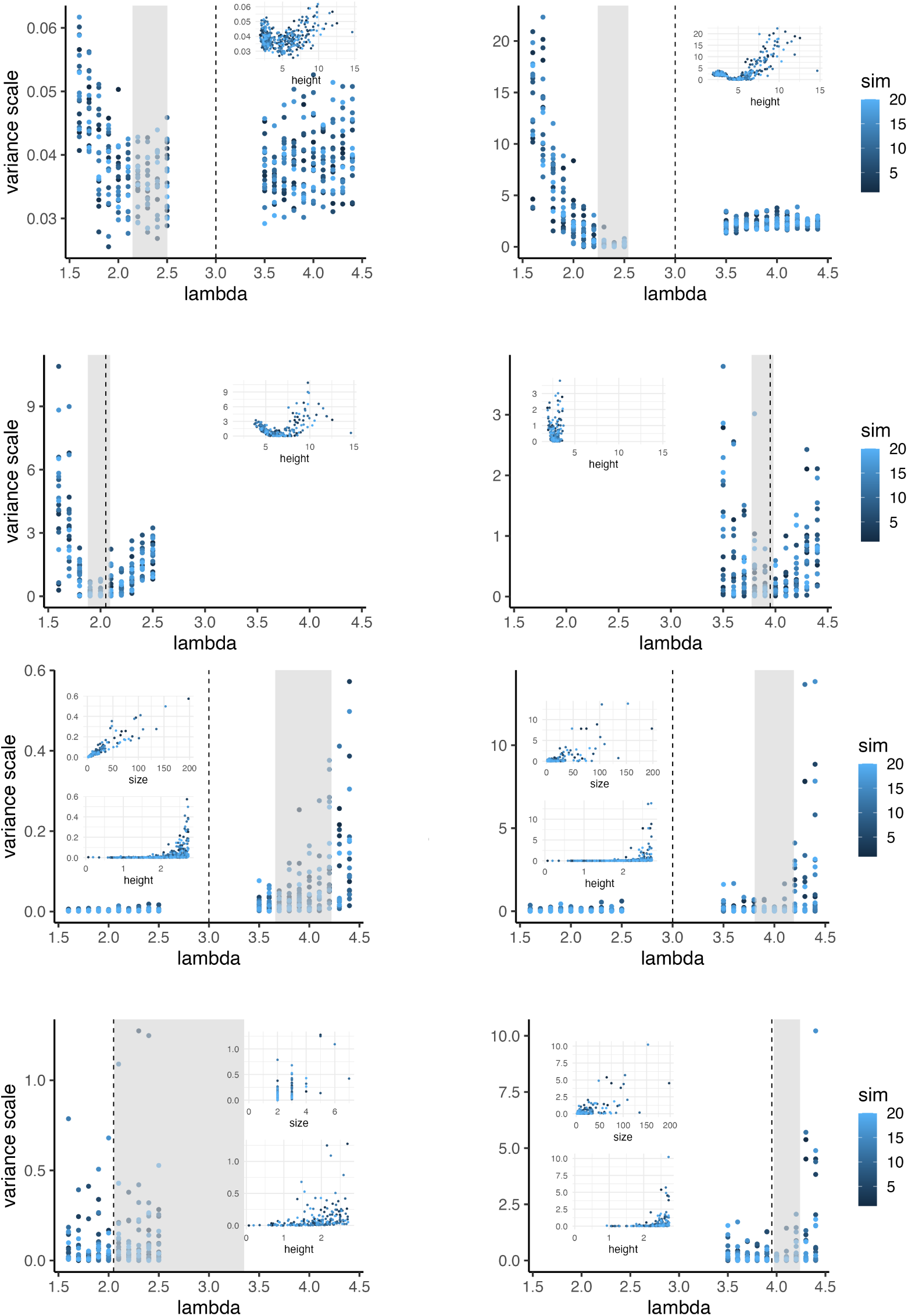
Drivers of cluster influence given independent non-identically distributed clusters: Scaled variance of the 20 cluster likelihoods for each of the 20 simulation replicates against the transmission rate used to simulate the respective cluster (size and height shown as insets). Replicates are indicated by colour (legend ’sim’); the dashed vertical line corresponds to the mean of the rate values used for simulation, the grey area to the range in which the means of the posterior distributions fall. The first two rows show results from simulation setting V, the last two rows from setting VI; rows one and three correspond to inferences from all 20 clusters jointly (left column inferring tree, evolutionary and transmission rate and right column inferring only the latter), rows two and four to inferences of the transmission rate based only on a subset of ten clusters (left column smallest ten rates, right column highest ten rates).

This is because clusters simulated under lower rates tend to have a higher influence on the shared rate. The effect is more pronounced for the influence on the transmission rate only (right column and Supplementary Figure 3), but also present for the influence on the tree and rate (left column). Clusters with high influence tend to have higher trees, suggesting that higher trees generate a more peaked log likelihood and are therefore more informative for the transmission rate than shorter trees (insets of Figure 3). In Supplementary Figure 4 we show that long branches are generally more informative against high rates than short branches are against low rates. In the second row of Figure 3 we shorten the range of the transmission rates used for simulation to [1.6*y^−^*^1^, 2.5*y^−^*^1^] and [3.5*y^−^*^1^, 4.4*y^−^*^1^], respectively, by combining only the ten low-rate (left column) and ten high-rate (right column) clusters in one analysis. In both cases, we find the posterior centre to be closer to the simulation mean, however, still slightly pulled downwards for most of the replicates. The insets of both plots illustrate how, as before, the higher influence correlates with higher trees, therefore lower simulation transmission rates. However, more pronounced than before, we see how the influence on the transmission rate is strongly driven by the distance of the simulation rate of the respective cluster to the shared estimate. Clusters that agree well with the shared rate have low individual influence on the estimate, while clusters that are less likely under the shared estimate are more influential. To illustrate how the local influence of the low- and high-rate clusters in the full analyses relates to the subset analyses in Figure 3 (second row), we calculated the grouped influence, described in Section 4.1 and plotted in Supplementary Figure 5. Despite the deletion of the low-rate group (Figure 3 right column) leading to a bigger absolute difference in mean posterior estimates to the full analysis, the grouped influence is indistinguishable from the high-rate clusters (Supplementary Figure 5 left column). When we divide the data into four groups, we see the lowest- or highest-rate group, respectively, to generally have the highest influence. The discrepancy observed for two groups can be the result of the different likelihood reweighting approach (local influence vs. case deletion) or of the different distance measure (Kullback-Leibler divergence vs. absolute difference of mean). Considering the latter, however, it would not be surprising: the Kullback-Leibler divergence takes into account the full probability distribution, not just the behaviour around the centre, and quantifies the expectation under the reweighted posterior of the logarithmic difference between the full and reweighted posterior (Kullback, 1959). In addition to the variance, we can consider the covariance of the cluster likelihoods to indicate the direction in which each cluster pulls the estimate. In Supplementary Figure 6 we colour the variances of Figure 3 by their respective covariance sign, i.e. the pull direction. In both the first and second row, the pull direction perfectly aligns with the position of the simulation rate of each cluster relative to the shared estimate.

##### Combined impact of rate, size and height (setting VI)

Finally, to include the effect of variable cluster sizes, we simulate birth-death-sampling trees with rates as above, allowed to grow until a maximum of 2.75*y*. Similarly as for the i.i.d. trees, the size has a strong impact on the influence of each cluster (Figure 3 row three and four). As expected, trees that were simulated under higher transmission rates tend to generate bigger and higher trees (insets). These also show the highest influence values, particularly when estimating both the transmission rate and the tree. As before, when only the influence on the transmission rate is considered (right column), we find big clusters with low influence. This can, for example, be caused by the impact of the true transmission rate, as clusters that agree well with the shared estimate have lower influence. Nevertheless, the posterior centre of the transmission rate is shifted to much higher values than in the previous simulation settings, with values ranging around 4.0*y^−^*^1^. To reduce the variability of the joined clusters, we again run the same analyses with only the 10 low-rate and 10 high-rate clusters, respectively (Figure 3 row four). For the low-rate group (left column), the strongest impact on the cluster influence seems to be mediated by the height of the tree (insets). However, for some of the simulation replicates the transmission rate is overestimated, as the centre of the posterior distribution is higher than the highest transmission rate used for simulation. As the cluster size in this setting ranges between 2 and 7 sequences, the overestimation of the rate can be the result of combining many likelihoods with very small log variance. For the high-rate group, we find a similar pattern as for the full data, with the shared estimate being pulled to values slightly higher than the simulation-mean by the high-rate trees. Nevertheless, when considering the influence by group in the full analysis, we cannot detect strong differences between the low-and high-rate clusters; only when separating the clusters into four groups (Supplementary Figure 5). In contrast to the previous simulations, the pull direction, coloured in Supplementary Figure 6, does not perfectly align with the relative position of the respective cluster to the shared estimate. However, this is particularly pronounced for the analysis of the low-rate group, suggesting that the small clusters mediate this effect. We, for example, find multiple clusters simulated under smaller rates to pull the shared estimate upward – which in turn can explain the overestimation.

Taken together, if clusters that were simulated under different transmission rates, are joined in one analysis, the less the cluster agrees with the shared estimate, the higher the influence on it is. This influence is additionally impacted by the size and persistence of the cluster, with bigger and longer persistent clusters often mediating higher influence. For the shared estimate this means that a complex interplay of the properties of the combined clusters relative to each other determines its value. This value then represents the probabilistically most plausible description of the combined clusters.

### 2.3 Transmission dynamics of SARS-CoV-2 clusters

To illustrate which insights can be practically gained through the assessment of cluster influence, we further inferred a shared reproductive number from pathogen genome sequences. More precisely, we analysed clusters of identical SARS-CoV-2 genomes sampled in Germany before the first nation-wide strict non-pharmaceutical interventions were implemented. We chose SARS-CoV-2, as most studies presenting shared Bayesian estimates so far focus on this virus. We restricted our analyses to sequences sampled in Germany before the first lockdown measures to obtain a data set of sensible size that allows the inference of one time-constant reproductive number. We focus on clusters of identical sequences, as these present the highest resolution of transmission chains that genome sequences can directly provide. Practically, this also means that the data for the phylogenetic likelihood is little informative and the analysis should be mainly driven by the sampling times through the birth-death-sampling likelihood.

As commonly described for fast spreading pathogens (Tsui et al., 2025), we find a power law-like distribution of cluster sizes, with many clusters consisting of less than five sequences and few of more than 30. Joining all clusters from which a phylogenetic tree can be constructed, that is those that contain at least two sequences, into one analysis, we inferred one shared, time-constant reproductive number. We estimate a ragged posterior density with a mean of 2.80 and a highest posterior density interval (HPDI) of [2.60, 2.91] (Figure 4 row 1 column 1). From the previous simulations, we expect this inference to be mainly driven by the biggest cluster, containing 290 sequences. Indeed, we find it to have the highest local influence (Figure 4 row 1 column 2). Other than expected, we also discover that the smallest clusters included in the analysis, containing only two sequences, have a high influence on the shared reproductive number. When grouped together, their influence even substantially exceeds the influence of the biggest cluster (Supplementary Figure 7). However, not all two-sequence clusters are associated with higher influence; the high-influence subset exclusively consists of clusters for which both sequences were sampled on the same day, such that the corresponding phylogenetic trees are ultrametric (Figure 4 row 1 column 2). Inspecting the MCMC chain of the parameters associated with these clusters, we see that progressively small tree heights are sampled, as they increase the birth-death-sampling likelihood. For the smallest heights, though, the likelihood becomes ragged and non-smooth, such that it jumps between three likelihood modes for all sampled values of the reproductive number (Supplementary Figure 8). For the two ultrametric clusters that are not associated with increased influence, one with two and one with three sequences, the sampled tree heights do not extend to such small values and the sampled likelihood behaves smoothly. As this is indicative of numerical instability due to the extremely small tree heights, the high influence is likely a numerical artefact potentially leading to biases in the shared estimate. We note, however, that we condition the birth-death-sampling probability density on the time of the first transmission event, that is the most recent common ancestor in the phylogenetic tree, rather than on the beginning of the process. In analyses using the latter conditioning this artefact might not be present for ultrametric two-tip trees. In addition, we did not observe similar behaviour in our simulations; this is unsurprising as under the simulation conditions the temporal resolution is higher such that sampled tips are very unlikely to fall on the exact same time point and clusters with truly ultrametric trees are therefore not included. Indeed, when excluding all ultrametric trees, the estimated posterior distribution of the reproductive number is unimodal, with a clear peak around the mean value 2.73 and the HPDI covering the interval [2.58, 2.89] (Figure 4 row 2 column 1). The assessment of the local influence for all clusters reveals that now the small clusters have little influence on the estimate, individually as well as grouped together (Supplementary Figure 7). Instead, the two biggest clusters have the highest influence, pulling the estimate in different directions (Figure 4 row 2 column 2). To determine in which absolute direction the clusters pull the estimate, we ran a third analysis excluding the biggest cluster (Figure 4 row 3). We find the resulting posterior distribution of the reproductive number to be broader and pulled downwards, with a median of 2.54 and HPDI of [2.31, 2.77]. This suggests that the biggest cluster with 290 sequences, as most other clusters, pulls the estimate to higher values while the second biggest with 49 sequences is more likely under lower reproductive numbers. This agrees well with previous phylodynamic analyses of the latter cluster, which support that after an initial superspreading event in the district of Heinsberg in mid-February 2020 the reproductive number fell to values below 1 during the time directly after the event until the end of April (Översti et al., 2025), when strict lockdown measures were already implemented regionally. Taken together, the assessment of the local influence of all joined clusters does not only reveal to what degree the clusters agree about the shared estimate, but can also highlight potentially biasing model and software artefacts.

**Figure 4:**
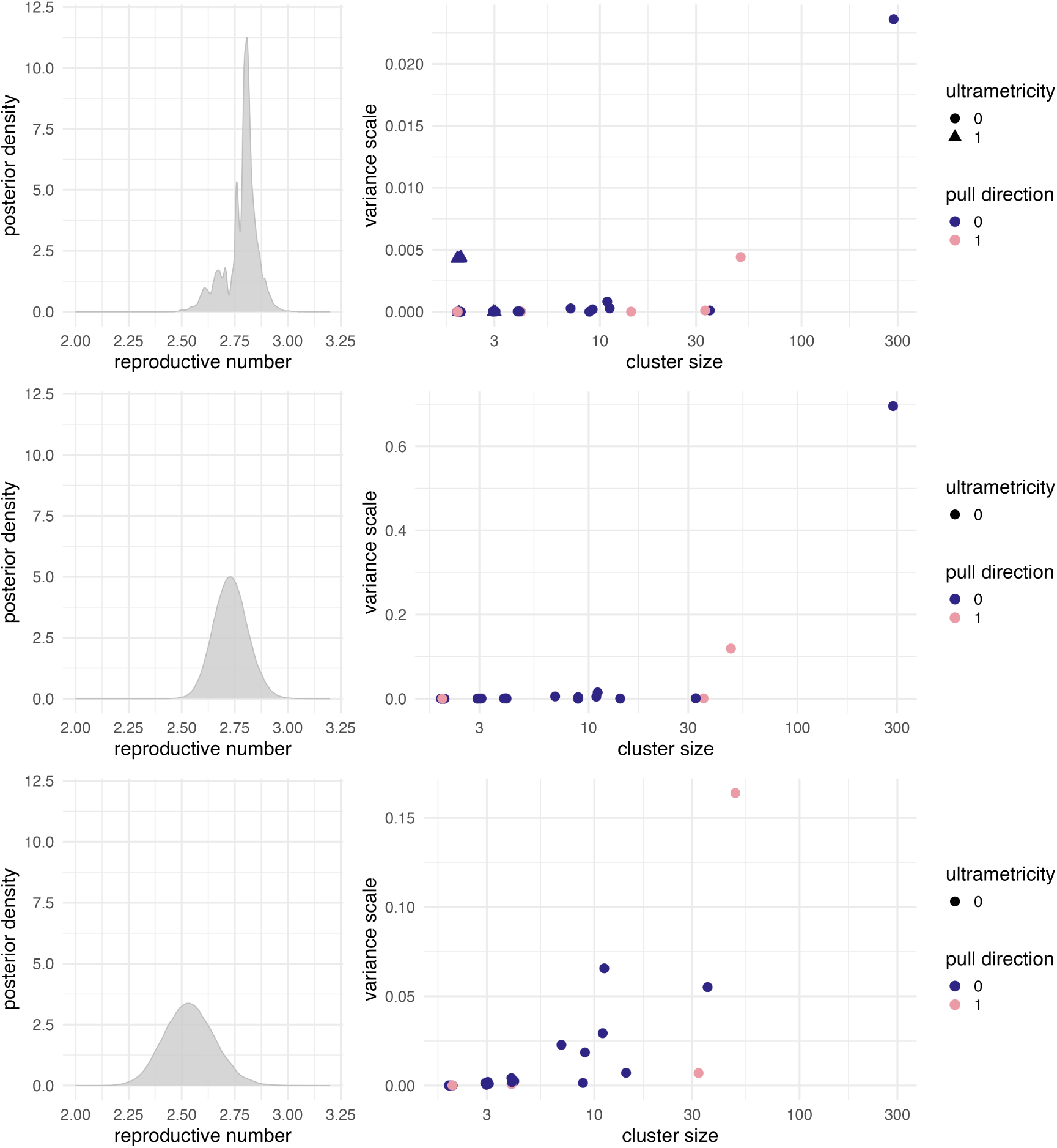
Shared estimate of the reproductive number from SARS-CoV-2 clusters and their individual influence: The left column depicts the sampled posterior distribution of the reproductive number. In the right column the variance of the logarithmic cluster likelihood marginalised for the transmission rate, i.e. the local influence on that rate only, is plotted against the size of the clusters. The colouring indicates the direction in which the cluster pulls the estimate, i.e. the sign of the covariance. Points that are shaped as triangles correspond to ultrametric clusters, that is clusters consisting only of sequences sampled on the same day, while circles to all others. The first row shows results from the analysis of all 59 clusters, the second of all 41 non-ultrametric trees and the third with all ultrametric trees as well as the biggest one excluded.

## 3 Discussion

The previous years have seen many applications of Bayesian phylodynamic models to the problem of estimating one shared parameter from multiple transmission clusters, assuming them to be conditionally independent realisations of the same process (Kühnert et al., 2018; Müller et al., 2020, 2021; Nadeau et al., 2023; Safina et al., 2022; M. R. Smith et al., 2021; Vasylyeva et al., 2019; Weber et al., 2023). As clusters of varying information content are joined together, it is important to understand to which extent the shared estimate is representative of them. In addition, Bayesian phylodynamic models are very complex structures that have been shown to be sensitive to various violations of model assumptions (Louca et al., 2021; Möoller et al., 2018; Översti et al., 2023; E. M. Volz & Frost, 2014). It is therefore particularly relevant to question and validate the inference results. To this end, we have presented a mathematical framework that can be used to easily quantify the information content that each cluster provides for the shared estimate, through the local influence of its likelihood functions. We have applied this framework to synthetic as well as real data to evaluate the formal drivers of influence, as considered here, and illustrated the practical insights that this framework allows. Focusing on birth-death-sampling models as tree generating processes, we find the shared estimate to be an average over all clusters that is relatively weighted by the certainty that each cluster provides about the parameter. Bigger sampling sizes and longer persistence of the cluster are generally associated with higher certainty and therefore higher influence on the joint analysis. The difference of the cluster-defining parameter to the shared estimate additionally mediates higher influence. All three characteristics strongly interact in determining the peakedness of its logarithmic cluster likelihood. The tree size, for example, can itself be informative about the birth-death-sampling parameters (MacPherson & Pennell, 2024). As this relative interplay is not trivial to predict, it is important to assess it on a case-by-case basis, for example through the local influence of each clusters presented herein.

Our results have important implications for the interpretation of shared estimates of the transmission rate in epidemiological contexts. Contrary to what is often assumed, the shared estimate does not correspond to a value that can be directly interpreted on the population-level but rather to an abstract value that is generated through the mathematical integration of uncertainties. Simulation setting II, which isolates the impact of the tree size, for example, demonstrates how the shared estimate is not equal to the cluster mean or median, as clusters of bigger sizes pull the estimate towards values that are more probable under their likelihoods. The shared estimate neither corresponds to the population mean or median, as illustrated by simulation setting V, in which clusters with longer persistence times have higher influence on the inference, given the same restrictions on the population size, purely due to them being stochastically more informative against specific value ranges. Instead, it will depend on the mathematical formulation of the inference problem, the specific setting and the relative interactions of all clusters included in the analysis, which epidemiological population-level entity, if any, the shared estimate best matches. Instead of estimating a shared rate, especially in the likely presence of rate heterogeneity between clusters, it is therefore often more sensible to employ models that can account for differences in the respective rates: hierarchical models can, for example, be used to constrain the rate of each cluster by a governing distribution (Edo-Matas et al., 2010; Taouk et al., 2024). Alternatively, individual rates can be inferred for each cluster and the resulting distributions merged in another post-inference Bayesian model. Comparisons of shared estimates of the reproductive number with quantifications of it from other models therefore have to be undertaken with care, as a direct comparison usually is not possible. As for the reproductive number, in particular, multiple definitions exist with slight differences in the population-level interpretation, the inter-comparability problem has already been discussed for other methods (Brockhaus et al., 2023).

Influence assessment can serve as a first step in better understanding in which way which parts of the analysed data set inform the inference, particularly because it is computationally undemanding to calculate for any MCMC-based sample from a posterior distribution. Thus, we recommend using it as a default diagnostic tool in joint Bayesian phylodynamic analyses. For other applications, like multi-level mediation models or regression analysis similar frameworks have allowed the identification of influential observations in specific model layers or data subtypes (Thomas et al., 2018; Věcěrováa et al., 2020). Our analyses of SARS-CoV-2 data demonstrate how employing it as a diagnostic tool does not only provide information about the epidemiological relevance of the clusters but can also hint towards computational artefacts.

It is, nevertheless, important to keep in mind what exactly our framework assesses: Influence is defined by the difference that a geometric reweighting of the cluster likelihoods causes on the posterior distribution, relative to the unweighted posterior. This local weighting of the cluster likelihood in a phylodynamic posterior distribution is not equivalent to a its complete deletion. Indeed, cluster-deleted analyses are therefore not directly comparable to local influence assessment, as we illustrate in Figure 4. In addition, there is no perfect way to measure the difference between two probability distributions. In our approach we follow Millar and Stewart, 2007 and Thomas et al., 2018 by calculating it as the Kullback-Leibler divergence, which can be understood as the relative lack of information provided by one distribution about the other. Importantly, it is not symmetric, meaning that the divergence between distribution A and B does not match the divergence between B and A (see Kullback, 1959 for a general derivation of the divergence score and McElreath, 2020 for a discussion of its use in Bayesian predictive accuracy). Symmetric measures of distance between distributions, as the Wasserstein distance, could alternatively be considered. In Supplementary Figures 1 and 2, we exemplify how both decisions, influence definition and difference measure, can impact the quantification of influence.

As for the empirical results that we provide using birth-death-sampling models as tree gener-ating processes in Bayesian phylodynamics, these have to be qualified by their specific contexts. Influence assessment is by definition highly relative and as such our decisions about the simulation settings will have a strong impact on our conclusions. In our simulations we opted to analyse a set of 20 clusters that varied in size between two and multiple hundred sequences and were generated by a birth rate between 2*y^−^*^1^ and 4*y^−^*^1^, a death rate of 1*y^−^*^1^ and a sampling rate of 0.01*y^−^*^1^. We chose these values for computational convenience, as multiple simulation replicates on a significantly bigger data set would take much more compute power, and due to their comparability to similar studies (Boskova et al., 2014). Different settings, however, might lead to different conclusions, although we expect the same statistical principle described above. Additionally, we only analysed trees generated by time-constant rates, while practical applications often involve much more complex models with the transmission rate, for example, changing tens to hundreds of times. As especially the influence of a single of multiple estimated parameters is based only on an approximation, it is likely to perform less good the more parameters are estimated. Nevertheless, in this study we opted for the simpler case to better isolate individual effects, like the numerical issues associated with ultrametric two-tip trees shown in Figure 4. As the basic influence measure applies for different models, we do expect similar patterns when other models, like coalescent-based prior distributions on the tree, are used: the influence of each observation will be driven by its degree of disagreement with the other observations as well as general factors that increase informativeness like sample size. In our case, sample size could easily be measured by tree size, i.e. tips in the tree, as only one underlying population is modelled. For multitype models, which combine several populations in one tree, sample size will be determined by a combination of the number of branches and nodes assigned to the respective types. In addition, if number and timing of type change events in the tree are not fixed but inferred, the sample size is not a constant property anymore but will change during the MCMC analysis. For inference problems, in which observations are not conditionally independent, further research is needed.

In summary, the presented method can serve as a convenient tool to evaluate inferences of shared parameters from multiple clusters. In this study, we focused on the inference of population dynamic parameters from a birth-death-sampling model, but the analytical framework can straightforwardly be extended to other parameters, like the evolutionary rate of the phylogenetic likelihood. Similarly, other inference settings can also be analysed: the main assumption of the method is conditional independence of the observations, which is also fulfilled by individual sites in an alignment under Felsenstein’s likelihood (Felsenstein, 1981). Extensions of influence assessment on groups of genomic sites could, for example, highlight those that drive our inference of evolutionary rates or those that might be in disagreement with the signal from other genomic positions, therefore pointing towards important evolutionary patterns.

## 4 Methods

### 4.1 Local influence in Bayesian phylodynamics

#### Local influence

We calculated the variance-covariance matrix of posterior samples of the logarithmic cluster likelihoods in the software environment R 4.4.2 (R Core Team, 2024) using the graphical user interface RStudio 2024.12.0+467. To evaluate the quality of the posterior samples we applied functions from the packages boa 1.1.8-2 (B. J. Smith, 2007) and coda v0.19-4.1 (Plummer et al., 2006), for parsing of simulated trees packages ape 5.8-1 (Paradis & Schliep, 2019) and phytools v2.4-4 (Revell, 2024) and for plotting of the results packages from the tidyverse collection v2.0.0 (Wickham et al., 2019).

#### Inference of multiple parameters

To approximate the influence of a single parameter in a BEAST2.7.7 (Bouckaert et al., 2019) analysis when inferring multiple parameters, as discussed in Section 2.1, we recalculated the log cluster likelihoods for all posterior samples of the parameter-of-interest while fixing all other inferred parameters to their median estimate. The posterior sample of phylogenetic trees we summarised by the maximum clade credibility (MCC) tree with median heights. The MCC trees were reconstructed with treeannotator, which is part of the BEAST2.7.7 (Bouckaert et al., 2019) distribution, while the cluster likelihoods were recalculated with the Log-FileIterator class, included in the package feast v10.4.0 (T. Vaughan, 2025).

### 4.2 Drivers of cluster influence

#### 4.2.1 Simulations of identically distributed clusters

##### Setting I: Impact of tree height

We simulated bifurcating trees distributed according to a birth-death-sampling process with birth rate *λ* = 2 per year (*y^−^*^1^), death rate *µ* = 1*y^−^*^1^ and sampling rate *ψ* = 0.01*y^−^*^1^ in the software BEAST2.7.7 (Bouckaert et al., 2019) with functionalities included in the package ReMaster v2.7.2 (T. G. Vaughan, 2024). To study the impact of the tree height on local influence, we controlled for the tree size by setting as simulation end condition 20 sampled tips. We generated a set of 20 trees for each of 20 simulation replicates. Along all trees we simulated sequences of 5000 nucleotides in length under the Jukes-Cantor-69 (JC69)(Jukes & Cantor, 1969) model with a rate of 0.002 substitutions per site per year (*ss^−^*^1^*y^−^*^1^). Using these nucleotide sequence alignments we set up two Bayesian inferences in BEAST 2.7.7 with the package BDSKY v1.5.1 (Stadler et al., 2012) for each replicate: i) estimating the tree, the transmission rate as well as the evolutionary rate and ii) estimating only the transmission rate. For both inferences, we fixed the remaining parameters to the true value, conditioned the birth-death-sampling process on the root node and set LN (0.0, 1.25^2^) as prior distribution on the transmission rate and *U* (0, 1) on the evolutionary rate, when applicable. We approximated the posterior distribution of the parameters-of-interest through MCMC-based sampling from the respective distributions, running the chain for 1.5 ∗ 10^7^ and 5 ∗ 10^5^ steps, sampling every 10000th and 1000th iteration, respectively. We further approximated the influence only on the transmission rate in inference i by recalculating the birth-death-sampling likelihood for each posterior sample of the transmission rate with the trees fixed to the respective MCC trees with median heights, as described in Section 2.1.

##### Setting II: Impact of tree size

To evaluate the isolated influence of the tree size on local influence, we controlled for the tree height while allowing for variation in the number of sampled tips. We thus simulated, in the same software as above, 20 replicates of 20 trees generated by a birth-death-sampling process with birth rate 2*y^−^*^1^, death rate 1*y^−^*^1^ and sampling rate 1.01*y^−^*^1^. To keep the tree height comparable between trees and replicates, we enforced a transmission event to happen at time 0.0*y* and 3.0*y* while setting a maximum simulation time of 3.0*y*. Again, we simulated 5000 nucleotide long sequences along each tree under the JC69 model with a rate of 0.002*ss^−^*^1^*y^−^*^1^ and used these sequences as input data in two Bayesian inference settings: i) estimating the tree, transmission rate as well as evolutionary rate and ii) estimating only the transmission rate.

##### Setting III: Combined impact of tree height and size

We studied the combined local influence of tree height and size by simulations of birth-death-processes with birth rate 2*y^−^*^1^, death rate 1*y^−^*^1^ and sampling rate 0.01*y^−^*^1^ and a maximum simulation time of 6.0*y*, conditioned on a minimum of two samples. We set up the simulation replicates and Bayesian analyses analogously to both previous sections.

#### 4.2.2 Simulations of non-identically distributed clusters

##### Setting IV: Impact of transmission rate

We evaluated the impact of differences in the tree generating process between jointly analysed clusters by simulating trees generated under different transmission rates. We note that in the following simulation settings the tree height is also not controlled for. First, we focused on the impact of a single transmission rate outlier: We simulated 19 trees from a birth-death-sampling process with birth rate 2*y^−^*^1^, death rate 1*y^−^*^1^ and sampling rate 0.01*y^−^*^1^ and one cluster with identical death and sampling rate but with a birth rate of 3*y^−^*^1^. We kept the size of all clusters constant by stopping the simulation when 20 samples were generated. The nucleotide sequences simulation and Bayesian inference were again set up as described above, notably with two inference settings: estimating i) the tree, evolutionary rate and transmission rate and ii) only the transmission rate.

##### Setting V: Combined impact of transmission rate and tree height

For the 20 replicates of 20 tree simulations, we set the death rate to 1*y^−^*^1^, the sampling rate to 0.01*y^−^*^1^ and the transmission rate to a value between 1.6*y^−^*^1^ and 2.5*y^−^*^1^, in steps of 0.1, for 10 trees as well as between 3.5*y^−^*^1^ and 4.4*y^−^*^1^, again in steps of 0.1 for further 10 trees for each replicate; such that each tree in one simulation replicate is characterised by a unique transmission rate. Keeping the tree size constant, we used as simulation end condition the generation of 20 samples. As for the previous four settings, we simulated nucleotide sequences along these trees, which we used as input in a Bayesian inference of i) the tree, transmission and evolutionary rate from all 20 clusters, ii) the transmission rate from all 20 clusters and iii) the transmission rate only from the the ten clusters simulated under lowest and highest rate, respectively.

##### Setting VI: Combined impact of transmission rate, tree height and size

As previously, we generated 20 trees for 20 replicates. Each tree was simulated from a process with death rate 1*y^−^*^1^, sampling rate 0.01*y^−^*^1^ and individual transmission rate between 1.6*y^−^*^1^ and 2.5*y^−^*^1^ and between 3.5*y^−^*^1^ and 4.4*y^−^*^1^. To also allow for variation in tree size, the simulations were stopped after 2.75*y* or upon depletion of the infected population. Sequence alignment simulation and Bayesian inferences were also set up as in simulation setting V.

### 4.3 Transmission dynamics of SARS-CoV-2 clusters

We collected all SARS-CoV-2 genomes from GISAID, the global data science initiative (Shu & McCauley, 2017), on January 19th 2025 that were sampled in Germany until March 22nd 2020, when the first nation-wide strict non-pharmaceutical interventions (’lockdown’) were started (Supplementary table 4). We restricted the search to include only complete, high coverage genomes with complete collection date information. This resulted in a data set of 826 SARS-CoV-2 genome sequences, which we aligned to the reference genome with GenBank-ID MN908947.3 (Wu et al., 2020) in mafft v7.490 (Katoh & Standley, 2013) with the alignment method chosen automatically and the alignment length kept constant. Sites that have been suggested not to contain phylogenetic signal (W, 2025), we replaced with an uninformative ’N’ in all sequences. We identified clusters as groups of identical genome sequences with minimal size of two in R 4.4.2 (R Core Team, 2024) with functionality from the packages ape (Paradis & Schliep, 2019) and tidyverse (Wickham et al., 2019), more precisely by calculating pairwise distances using the function dist.dna without speci-fication of a substition model and pairwise deletion set to false to avoid sequences being added to multiple clusters. This left us with 582 sequences in 59 clusters of a minimal cardinality of two, the biggest of which consisted of 290 sequences while most others contained less than five. We next set up a Bayesian inference in the software BEAST2.7.7 (Bouckaert et al., 2019) of one shared effective reproductive number *R_e_* from all clusters together with their individual phylodynamic trees. For this, we defined independent tree likelihoods for all clusters, calculated under a strict clock model with fixed rate 8 ∗ 10*^−^*^4^*ss^−^*^1^*y^−^*^1^ and the Hasegawa-Kishino-Yano-85 (HKY85) (Hasegawa et al., 1985) substitution model with estimated base frequencies and transition/transversion ratio. The prior distribution for each tree we specified to be the birth-death-skyline model from the package BDSKY v1.5.1 (Stadler et al., 2012) with the rate to become non-infectious *δ* fixed to 36.5 *y^−^*^1^ while the reproductive number *R_e_* and sampling proportion *s* were estimated and shared between all trees. We used the epidemiological, in contrast to the rate parameterisation in the simulation analyses, as its quantities can be directly interpreted while arising through the simple reparameterisation: 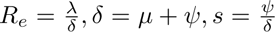, with *λ, µ, ψ* being the birth/transmission, death and sampling rate, respectively. We finalised the Bayesian model by defining prior probability distributions for all hyperparameters: the reproductive number was constrained by a LN (0.0, 1.25^2^) distribution, the sampling proportion by *U* (0.0, 0.1), the base frequencies by a standard uniform distribution and the transition/transversion ratio by LN (1.0, 1.25^2^). We sampled from the posterior distribution via a MCMC algorithm with 5 ∗ 10^8^ steps, storing every 10^4^th step for all parameters except the phylodynamic trees, which were stored every 7.5 ∗ 10^4^th step. We assessed convergence of the chain in R with the package coda (Plummer et al., 2006), running chains for which any estimated parameter did not yet have an effective sample size above 150 for another 2.5 ∗ 10^8^ steps. In addition, we ran two independent chains with different seeds of the random number generator and validated that both converged to similar values. To approximate the influence on a single parameter, we proceeded as described above. We further include two analyses with slightly different data sets. First, with all clusters that contain only sequences sampled on the same day removed, second with the biggest cluster additionally excluded. For both data sets we set up the Bayesian inference as described for the full data set and proceeded with the influence assessment analogously.

## Data Availability

The analyses of SARS-CoV-2 data are based on 826 sequences and their associated metadata available on GISAID up to January 19th 2025, via gisaid.org/EPI_SET_250119mc and under doi.org/10.55876/gis8.250119mc. Scripts, input files and the raw data shown in figures of the main text are published as a data set under https://doi.org/10.17617/3.DNLGXJ.

https://gisaid.org/EPI_SET_250119mc

## Acknowledgements

We gratefully acknowledge all data contributors, i.e., the Authors and their Originating laborato-ries responsible for obtaining the specimens, and their Submitting laboratories for generating the genetic sequence and metadata and sharing via the GISAID Initiative, on which this research is based.

## 5 Data availability

The analyses of SARS-CoV-2 data presented in Section 2.3 are based on 826 sequences and their associated metadata available on GISAID up to January 19th 2025, via gisaid.org/EPLSET250119mc and under doi.org/10.55876/gis8.250119mc. Scripts, input files and the raw data shown in figures of the main text are published as a data set under https://doi.org/10.17617/3.DNLGXJ.

## 6 Author contributions

Conceptualization: AW, RB, SD

Data curation: AW

Formal analysis: AW

Investigation: AW

Methodology: AW, RB, SD

Software: AW

Supervision: AW, SD

Validation: AW, RB, SD

Visualization: AW, RB

Writing – original draft: AW, RB

Writing – review & editing: AW, RB, SD

