## Supplementary Material for "Identification of transmission clusters with high influence on shared parameter estimates in Bayesian phylodynamics"

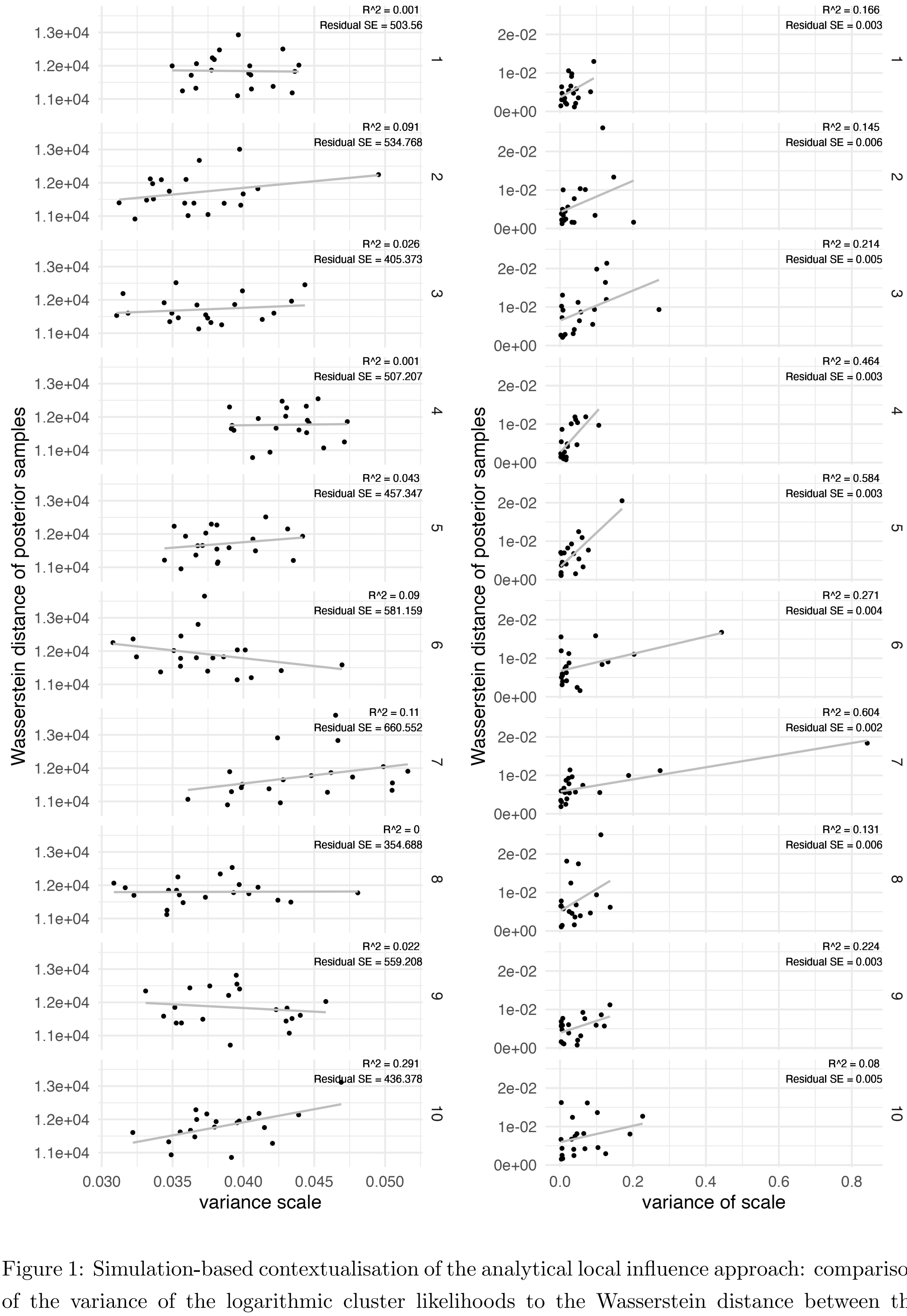


Figure 1: Simulation-based contextualisation of the analytical local influence approach: comparison of the variance of the logarithmic cluster likelihoods to the Wasserstein distance between the full and cluster-deleted posterior distribution for identically distributed clusters. Simulations are described in section 3.2.1 of the main text (simulation setting I), inferences are set up to estimate the tree, transmission rate and evolutionary rate. The plots in the left panel compare the variances of the cluster likelihoods to the Wasserstein distances between samples from the full and cluster-deleted posterior distribution, plots in the right panel compare the variances of the logarithmic cluster likelihoods, marginalised for the influence on the transmission rate, as described in section 3.1, to the Wasserstein distances between the full and cluster-deleted posterior samples of the transmission rate. Plots in each row show the result of one of ten simulation replicates.

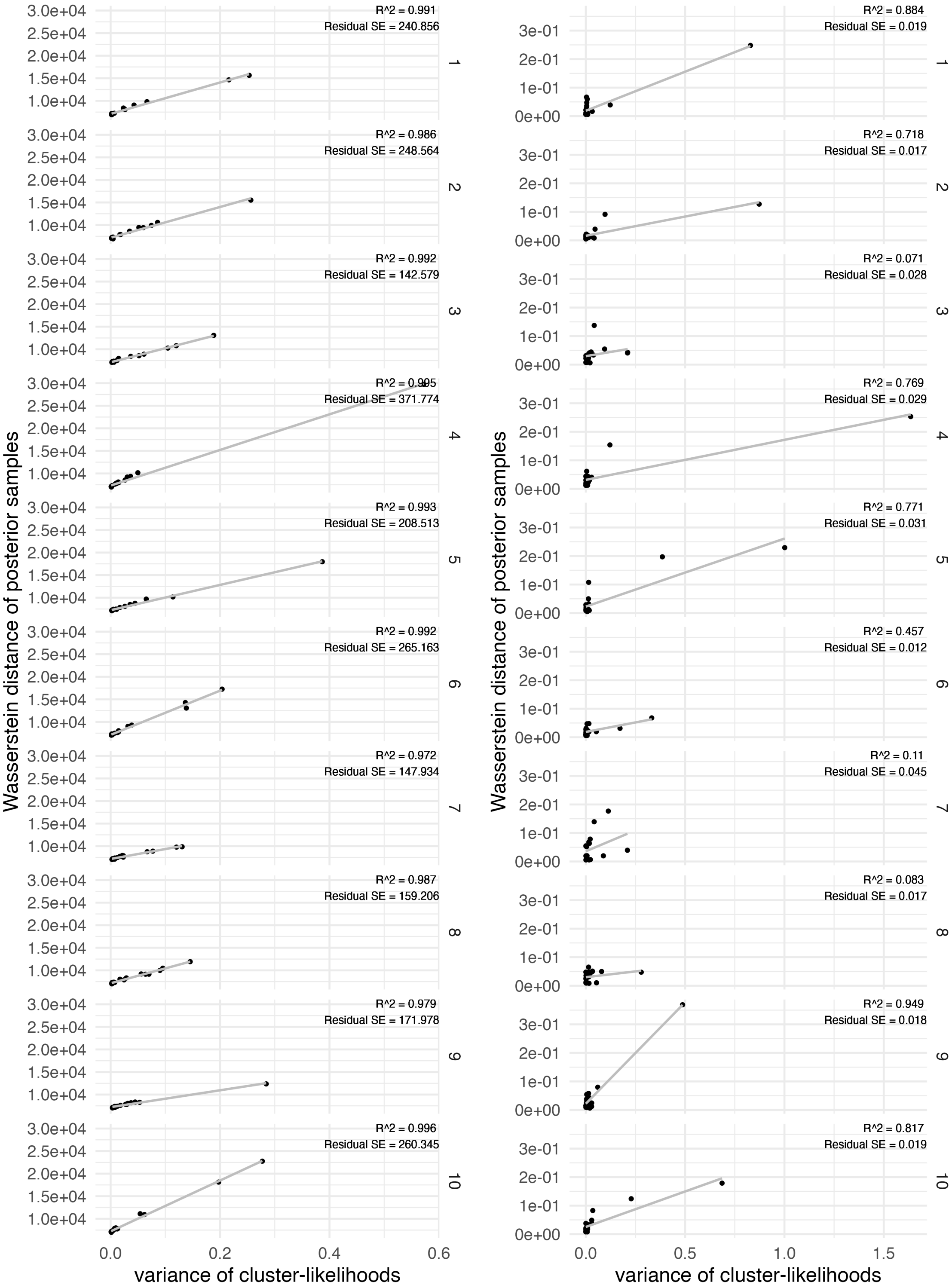


Figure 2: Simulation-based contextualisation of the analytical local influence approach: comparison of the variance of the logarithmic cluster likelihoods to the Wasserstein distance between the full and cluster-deleted posterior distribution for non-identically distributed clusters. Simulations are described in section 3.2.2 of the main text, inferences are set up to estimate the tree, transmission rate and evolutionary rate. The plots in the left panel compare the variance of the logarithmic cluster likelihood to the Wasserstein distance between samples from the actual posterior distribution, the plots in the right panel compare the variance, marginalised for the influence on the transmission rate as described in section 3.1, to the distance between the posterior samples of the transmission rate. Plots in each row show the result of one of ten simulation replicates.

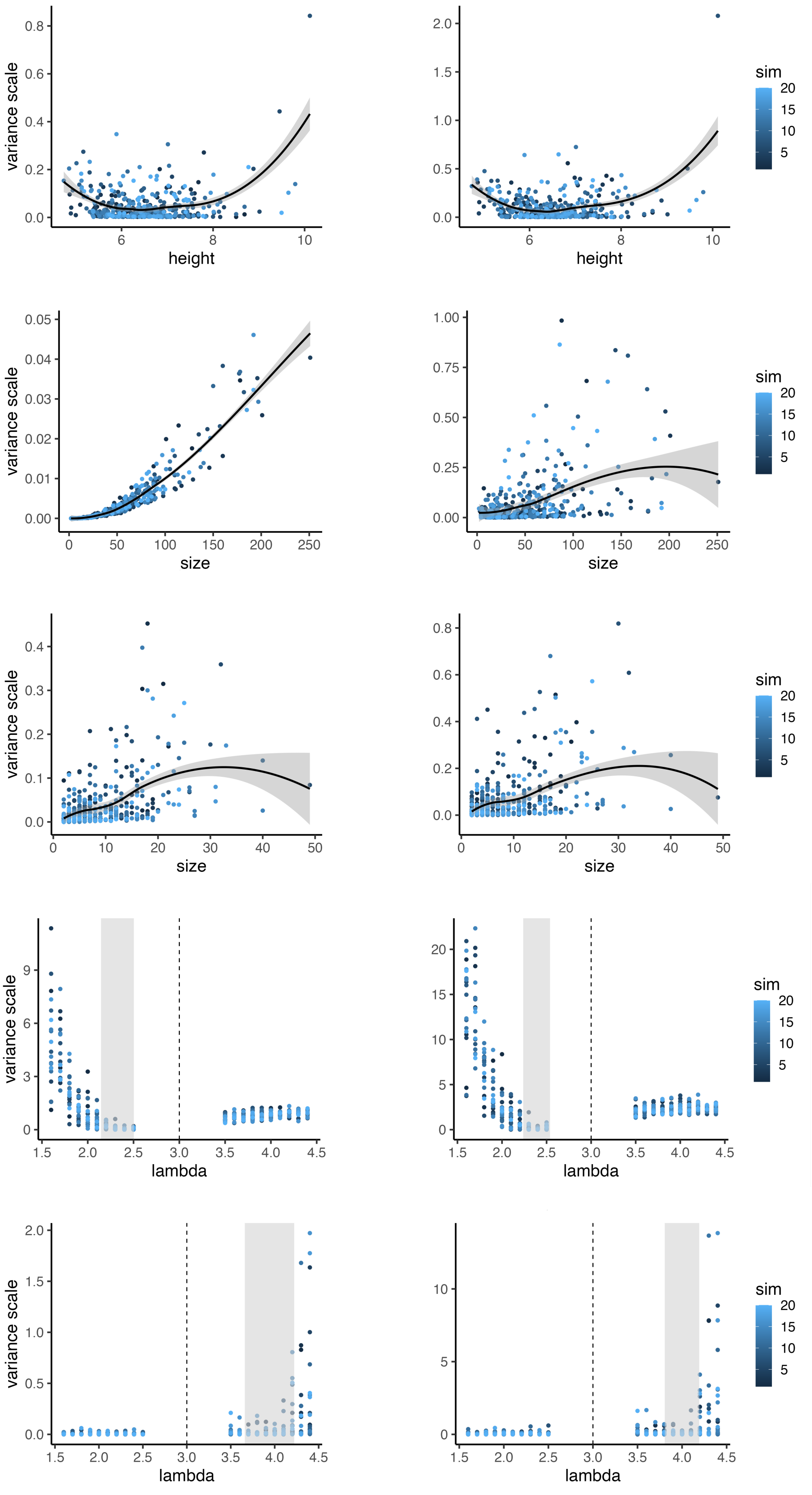


Figure 3: Simulation-based contextualisation of the approximation of the influence of a subset of the estimated parameters: scaled variance of the logarithmic cluster likelihoods against the respective parameter (transmission rate). Plots in the left panel show the scaled variances of the logarithmic cluster likelihoods that were recalculated from the posterior samples described in Sections 3.2.1 and 3.2.2, fixing all the parameters to the median estimate except for the transmission rate; plots in the right panel the results for inferences of only the transmission rate (i.e. all other parameters are fixed to the truth during the MCMC), as shown in Figures 1 and 3.

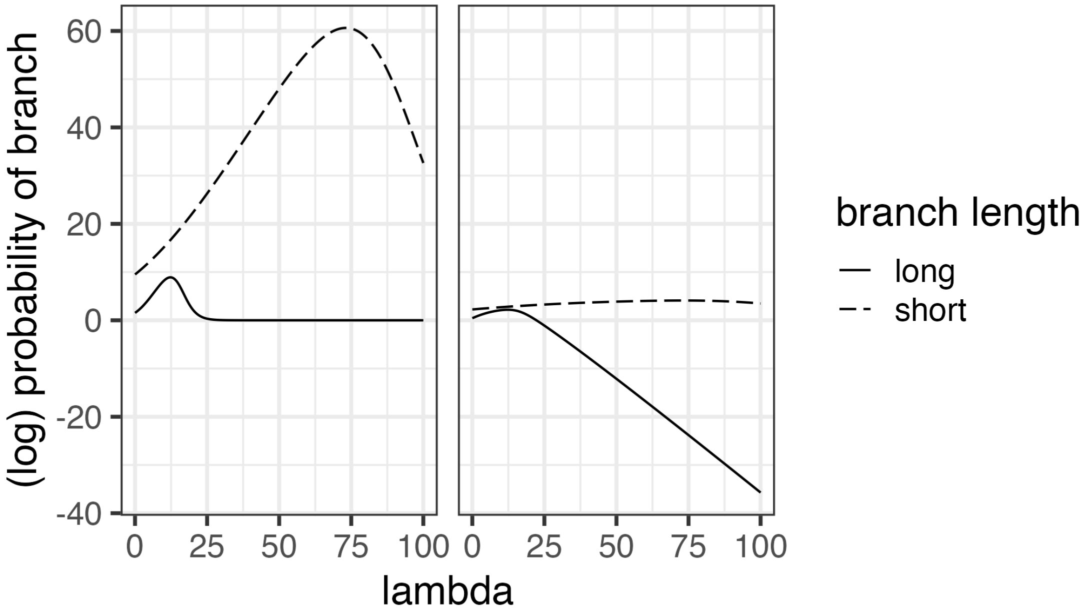


Figure 4: Informativeness of different branch lengths: birth-death-sampling likelihood for a short and long branch against the transmission rate. In the left panel the probability of a one-tip birth-death-sampling tree is plotted against the transmission rate given a short (dashed line) and long (solid line) tree/branch. In the right panel the corresponding logarithmic probability is shown. The birth-death-likelihood is calculated in BEAST2, the height of the branch is adjusted using the "origin" parameter.

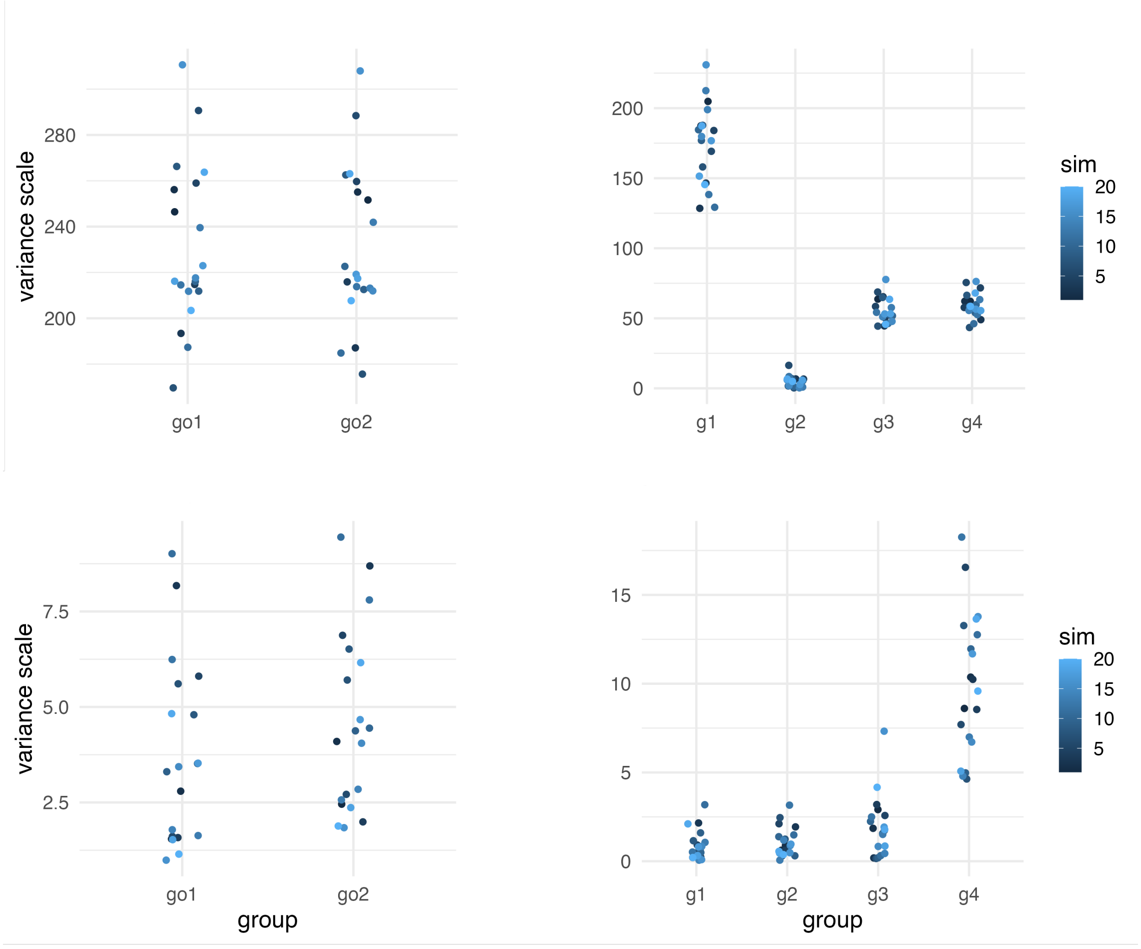


Figure 5: Influence of groups of simulated clusters: scaled variance of the logarithmic cluster likelihood against different groupings of the simulated clusters described in section 3.2.2. Plots in the left panel group clusters 1-10 and 11-20 together, in the right panel clusters 1-5, 6-10, 11-15 and 16-20. The first row is based on simulation setting V (fixed cluster size), the second row on simulation setting VI (maximum simulation time of 2.75y).

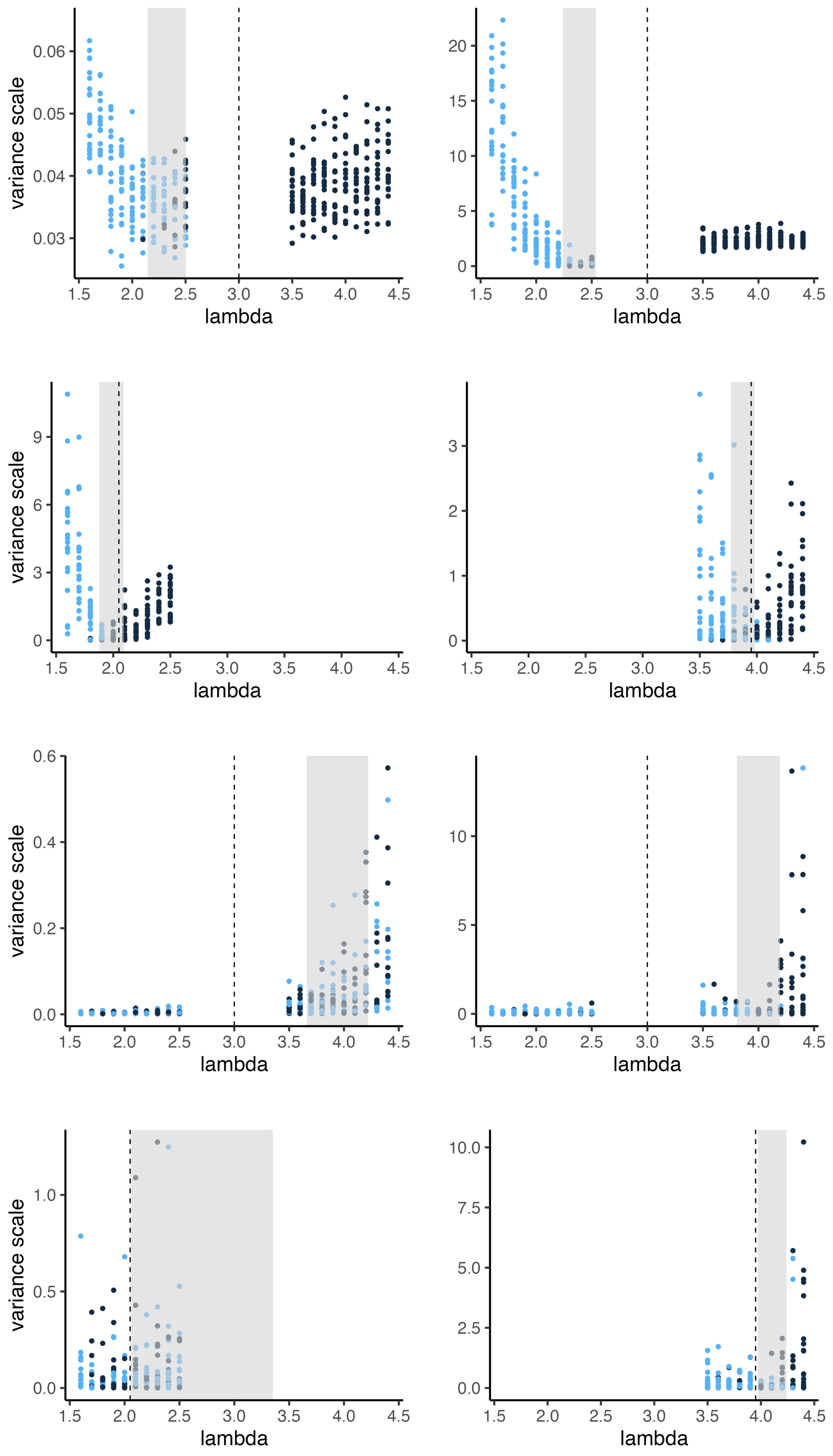


Figure 6: Direction in which each cluster pulls the posterior distribution: Figure 3 from the main text replotted, such that the colours of the points do not correspond to simulation replicate but to the relative direction in which the respective cluster pulled the full posterior distribution. The y-axis shows the scaled variance of the cluster likelihoods, the x-axis the transmission rate used for simulation of the respective cluster, the dashed line the mean of the rates used for simulation, the grey area the the interval that is covered by the means of the posterior distributions of the transmission rate and each point to one of 20 (first and third row) or 10 (second and fourth row) cluster in one of 20 simulation replicates. The relative pull direction is determined by the sign of the covariances.

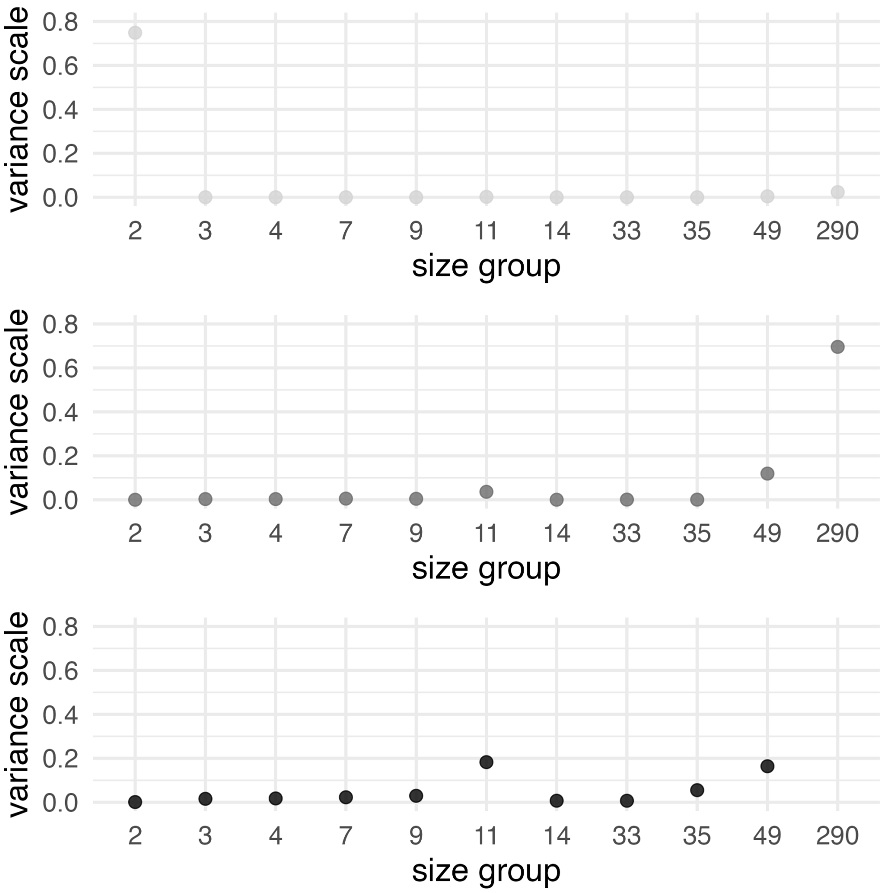


Figure 7: Influence of SARS-CoV-2 clusters grouped by size: Scaled variance of the cluster groups against the group for all three analyses presented in the main text. Clusters with identical sizes are grouped together. The first column shows the results from the full analysis, the second excluding all ultrametric trees, the third excluding all ultrametric trees and the biggest cluster.

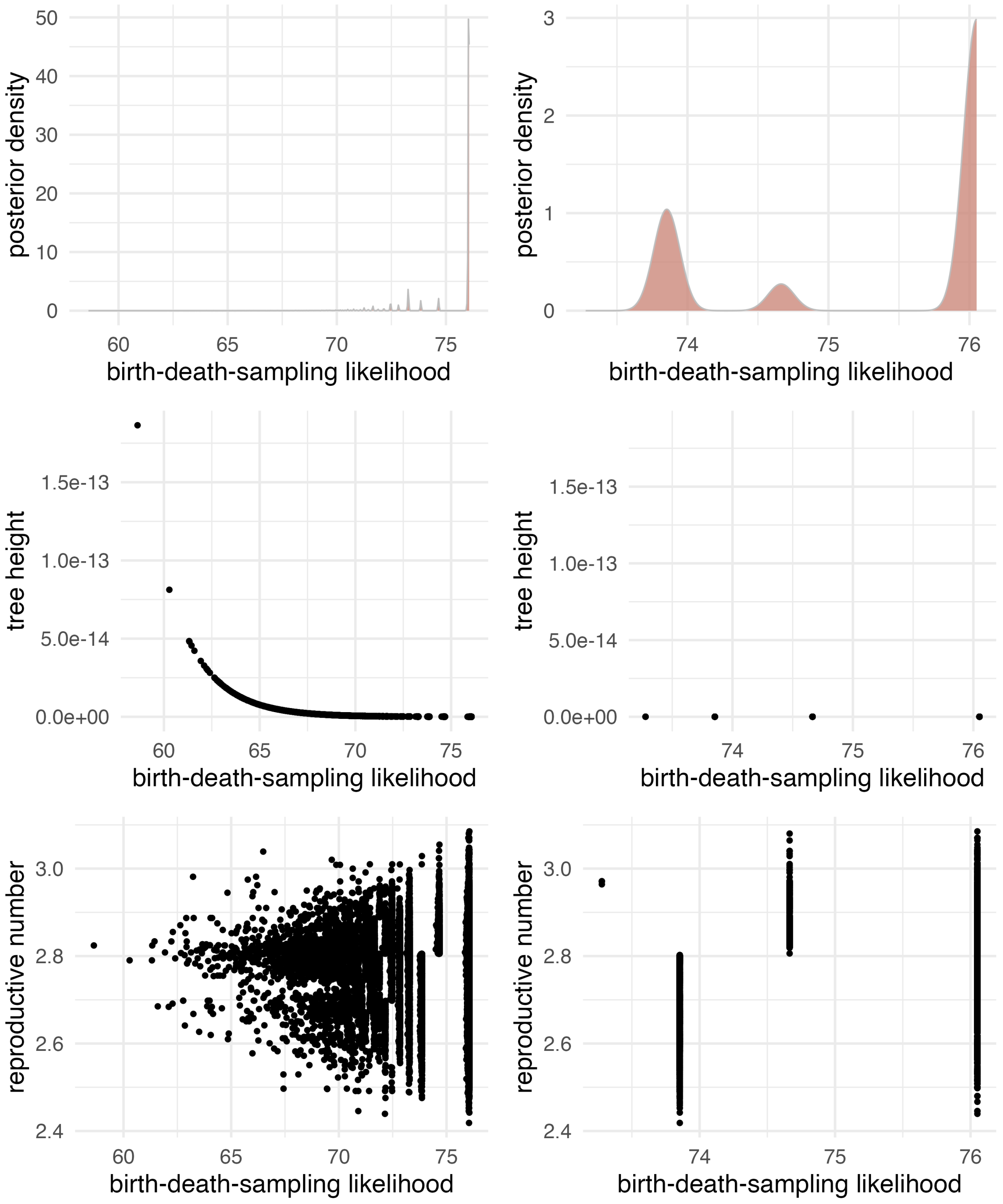


Figure 8: Behaviour of one ultrametric 2-sequence SARS-CoV-2 cluster: posterior density, tree height and reproductive number against the birth-death-sampling likelihood sampled during the MCMC approximation of the shared posterior distribution. The left column corresponds to the full analysis, the right column to the recalculation of the birth-death-sampling likelihood for all samples of the reproductive number with the remaining estimated parameters fixed to their median value. All other ultrametric 2-sequence behave similarly, except one for which the tree height is sampled to be around $0.02y$. The only ultrametric tree with more sequences, that is three sequences, neither shares this behaviour.

| rep. | I — all | I — lambda | II — all | II — lambda | III — all | III — lambda |
|---|---|---|---|---|---|---|
| 1 | 1.97 [1.91, 2.04] | 1.98 [1.94, 2.03] | 2.05 [1.93, 2.18] | 2.06 [1.95, 2.17] | 1.97 [1.86, 2.08] | 1.99 [1.91, 2.08] |
| 2 | 1.95 [1.89, 2.02] | 1.99 [1.95, 2.04] | 2.04 [1.91, 2.18] | 2.01 [1.88, 2.13] | 2.01 [1.92, 2.11] | 2.08 [2.00, 2.16] |
| 3 | 2.03 [1.96, 2.10] | 2.02 [1.97, 2.07] | 2.01 [1.92, 2.11] | 2.02 [1.92, 2.11] | 2.03 [1.92, 2.14] | 2.03 [1.94, 2.12] |
| 4 | 2.01 [1.94, 2.06] | 2.01 [1.97, 2.06] | 2.09 [1.99, 2.20] | 2.06 [1.96, 2.16] | 2.07 [1.95, 2.20] | 2.00 [1.92, 2.10] |
| 5 | 2.04 [1.97, 2.10] | 2.04 [1.99, 2.09] | 2.11 [2.01, 2.23] | 2.08 [1.98, 2.21] | 1.97 [1.83, 2.11] | 2.04 [1.95, 2.15] |
| 6 | 1.98 [1.92, 2.06] | 1.94 [1.89, 1.99] | 2.04 [1.93, 2.15] | 2.07 [1.95, 2.17] | 2.05 [1.97, 2.15] | 2.06 [1.99, 2.13] |
| 7 | 1.99 [1.93, 2.05] | 1.99 [1.95, 2.04] | 2.13 [2.01, 2.26] | 2.11 [2.00, 2.22] | 2.06 [1.96, 2.18] | 2.08 [1.99, 2.15] |
| 8 | 2.00 [1.94, 2.07] | 1.99 [1.95, 2.04] | 2.03 [1.92, 2.15] | 2.05 [1.95, 2.16] | 2.07 [1.96, 2.18] | 2.06 [1.97, 2.13] |
| 9 | 1.97 [1.91, 2.03] | 1.96 [1.91, 2.01] | 2.08 [1.97, 2.21] | 2.08 [1.98, 2.19] | 1.92 [1.81, 2.06] | 1.95 [1.86, 2.05] |
| 10 | 2.00 [1.95, 2.07] | 2.00 [1.95, 2.04] | 1.95 [1.83, 2.06] | 1.96 [1.85, 2.09] | 2.04 [1.94, 2.13] | 2.06 [1.98, 2.14] |
| 11 | 2.01 [1.95, 2.07] | 2.00 [1.95, 2.05] | 2.07 [1.95, 2.18] | 2.07 [1.96, 2.19] | 2.07 [1.98, 2.17] | 2.06 [1.97, 2.13] |
| 12 | 1.95 [1.89, 2.02] | 1.95 [1.90, 2.00] | 2.10 [2.00, 2.22] | 2.05 [1.98, 2.16] | 2.06 [1.95, 2.18] | 2.05 [1.97, 2.13] |
| 13 | 2.03 [1.97, 2.09] | 2.04 [1.99, 2.08] | 2.05 [1.92, 2.16] | 2.06 [1.93, 2.16] | 2.04 [1.93, 2.16] | 2.07 [1.99, 2.16] |
| 14 | 2.02 [1.96, 2.09] | 2.02 [1.97, 2.06] | 2.09 [1.98, 2.20] | 2.10 [1.99, 2.20] | 2.07 [1.98, 2.14] | 2.04 [1.98, 2.10] |
| 15 | 2.01 [1.93, 2.07] | 2.00 [1.96, 2.05] | 2.04 [1.93, 2.17] | 2.05 [1.95, 2.15] | 2.10 [2.00, 2.20] | 2.06 [1.98, 2.13] |
| 16 | 1.97 [1.91, 2.03] | 1.98 [1.93, 2.02] | 2.10 [1.98, 2.22] | 2.05 [1.94, 2.17] | 2.12 [2.00, 2.24] | 2.07 [1.99, 2.16] |
| 17 | 1.97 [1.90, 2.03] | 1.99 [1.94, 2.05] | 2.00 [1.87, 2.12] | 1.98 [1.87, 2.11] | 2.08 [1.99, 2.17] | 2.07 [2.01, 2.14] |
| 18 | 2.01 [1.95, 2.07] | 1.99 [1.95, 2.04] | 2.01 [1.90, 2.12] | 1.99 [1.89, 2.09] | 1.99 [1.87, 2.10] | 1.99 [1.92, 2.07] |
| 19 | 1.98 [1.91, 2.04] | 1.99 [1.94, 2.03] | 2.05 [1.93, 2.19] | 2.08 [1.96, 2.20] | 1.97 [1.87, 2.07] | 2.03 [1.96, 2.11] |
| 20 | 2.01 [1.95, 2.07] | 1.99 [1.94, 2.04] | 2.11 [2.00, 2.23] | 2.12 [2.02, 2.22] | 2.05 [1.96, 2.16] | 2.05 [1.98, 2.12] |

Table 1: Transmission rate estimates for all 20 simulation replicates of simulation settings I to III (identically distributed clusters) and their respective inference settings ('all': estimation of tree, evolutionary rate and transmission rate, 'lambda': estimation of transmission rate only): Mean of the posterior sample and 95% highest posterior density interval in square brackets.

| rep. | IV — all | IV — lambda | V — all | V — lambda | VI — all | VI — lambda |
|---|---|---|---|---|---|---|
| 1 | 1.98 [1.91, 2.04] | 2.01 [1.96, 2.06] | 2.38 [2.27, 2.47] | 2.42 [2.35, 2.50] | 4.03 [3.72, 4.34] | 3.95 [3.81, 4.08] |
| 2 | 2.00 [1.93, 2.06] | 1.99 [1.94, 2.04] | 2.23 [2.15, 2.33] | 2.24 [2.17, 2.30] | 4.06 [3.73, 4.36] | 4.05 [3.91, 4.18] |
| 3 | 2.06 [2.00, 2.12] | 2.03 [1.98, 2.08] | 2.15 [2.05, 2.23] | 2.33 [2.27, 2.42] | 4.01 [3.72, 4.35] | 4.00 [3.84, 4.14] |
| 4 | 1.98 [1.92, 2.05] | 1.99 [1.94, 2.04] | 2.43 [2.32, 2.53] | 2.47 [2.40, 2.55] | 4.04 [3.80, 4.29] | 4.14 [4.03, 4.26] |
| 5 | 2.03 [1.97, 2.10] | 2.02 [1.97, 2.07] | 2.49 [2.39, 2.60] | 2.54 [2.46, 2.62] | 4.22 [3.93, 4.51] | 4.05 [3.88, 4.17] |
| 6 | 2.02 [1.95, 2.08] | 2.00 [1.95, 2.05] | 2.29 [2.19, 2.40] | 2.41 [2.35, 2.49] | 4.11 [3.82, 4.38] | 4.10 [3.98, 4.23] |
| 7 | 2.04 [1.97, 2.10] | 2.04 [2.00, 2.09] | 2.29 [2.20, 2.38] | 2.36 [2.27, 2.43] | 3.73 [3.36, 4.14] | 3.81 [3.62, 3.98] |
| 8 | 2.03 [1.96, 2.09] | 2.01 [1.96, 2.06] | 2.30 [2.20, 2.39] | 2.33 [2.25, 2.40] | 4.18 [3.84, 4.53] | 3.98 [3.82, 4.12] |
| 9 | 1.95 [1.89, 2.02] | 1.97 [1.92, 2.02] | 2.42 [2.31, 2.51] | 2.40 [2.33, 2.48] | 3.67 [3.30, 4.04] | 3.86 [3.68, 4.06] |
| 10 | 2.02 [1.96, 2.08] | 1.99 [1.95, 2.05] | 2.35 [2.25, 2.44] | 2.37 [2.29, 2.44] | 4.01 [3.76, 4.23] | 4.15 [4.03, 4.27] |
| 11 | 1.99 [1.93, 2.06] | 2.04 [1.99, 2.09] | 2.32 [2.22, 2.41] | 2.39 [2.31, 2.47] | 4.02 [3.62, 4.42] | 3.93 [3.79, 4.12] |
| 12 | 2.03 [1.97, 2.10] | 2.00 [1.96, 2.05] | 2.50 [2.40, 2.61] | 2.49 [2.42, 2.57] | 4.11 [3.78, 4.45] | 4.09 [3.94, 4.25] |
| 13 | 1.96 [1.89, 2.02] | 1.96 [1.91, 2.01] | 2.35 [2.24, 2.45] | 2.41 [2.33, 2.47] | 3.93 [3.71, 4.15] | 4.10 [3.99, 4.22] |
| 14 | 1.96 [1.90, 2.03] | 1.96 [1.91, 2.00] | 2.31 [2.21, 2.40] | 2.37 [2.29, 2.44] | 4.07 [3.70, 4.38] | 4.06 [3.92, 4.21] |
| 15 | 2.03 [1.97, 2.10] | 2.02 [1.97, 2.06] | 2.32 [2.22, 2.42] | 2.40 [2.33, 2.48] | 3.95 [3.63, 4.23] | 3.90 [3.73, 4.03] |
| 16 | 1.96 [1.90, 2.02] | 1.96 [1.91, 2.00] | 2.44 [2.32, 2.54] | 2.51 [2.43, 2.58] | 4.05 [3.80, 4.31] | 4.20 [4.09, 4.34] |
| 17 | 1.99 [1.93, 2.05] | 2.00 [1.96, 2.06] | 2.36 [2.27, 2.45] | 2.41 [2.34, 2.48] | 3.97 [3.54, 4.35] | 4.06 [3.87, 4.26] |
| 18 | 2.01 [1.95, 2.08] | 2.03 [1.98, 2.08] | 2.32 [2.22, 2.42] | 2.34 [2.26, 2.41] | 3.98 [3.74, 4.23] | 4.12 [4.00, 4.24] |
| 19 | 1.99 [1.92, 2.05] | 2.00 [1.94, 2.05] | 2.34 [2.24, 2.43] | 2.34 [2.26, 2.40] | 3.71 [3.40, 4.04] | 3.82 [3.67, 3.99] |
| 20 | 2.05 [1.98, 2.11] | 2.02 [1.97, 2.06] | 2.45 [2.34, 2.54] | 2.48 [2.41, 2.56] | 4.09 [3.86, 4.29] | 4.13 [4.02, 4.22] |

Table 2: Transmission rate estimates for all 20 simulation replicates of simulation settings IV to VI (20 non-identically distributed clusters) and their respective inference settings ('all': estimation of tree, evolutionary rate and transmission rate, 'lambda': estimation of transmission rate only): Mean of the posterior sample and 95% highest posterior density interval in square brackets.

| rep. | IV — 1 to 10 | IV — 11 to 20 | V — 1 to 10 | V — 11 to 20 |
|---|---|---|---|---|
| 1 | 1.99 [1.92, 2.06] | 3.97 [3.82, 4.12] | 2.63 [2.00, 3.28] | 4.01 [3.86, 4.15] |
| 2 | 1.88 [1.81, 1.94] | 3.90 [3.74, 4.04] | 2.76 [2.11, 3.30] | 4.14 [3.98, 4.28] |
| 3 | 1.91 [1.85, 1.99] | 3.83 [3.70, 3.98] | 2.74 [2.32, 3.14] | 4.16 [4.02, 4.35] |
| 4 | 2.04 [1.97, 2.11] | 3.93 [3.78, 4.07] | 2.68 [1.98, 3.39] | 4.19 [4.07, 4.31] |
| 5 | 2.09 [2.02, 2.17] | 3.96 [3.83, 4.11] | 2.69 [2.15, 3.15] | 4.18 [4.02, 4.34] |
| 6 | 1.98 [1.91, 2.06] | 3.96 [3.81, 4.09] | 2.88 [2.33, 3.42] | 4.17 [4.04, 4.28] |
| 7 | 1.94 [1.87, 2.01] | 3.81 [3.65, 3.95] | 2.98 [2.53, 3.38] | 3.97 [3.78, 4.17] |
| 8 | 1.92 [1.86, 1.99] | 3.95 [3.83, 4.11] | 2.56 [2.09, 3.14] | 4.10 [3.97, 4.26] |
| 9 | 1.95 [1.87, 2.02] | 3.94 [3.81, 4.08] | 2.81 [2.25, 3.30] | 4.00 [3.82, 4.17] |
| 10 | 1.95 [1.87, 2.02] | 3.90 [3.76, 4.04] | 2.86 [2.40, 3.33] | 4.23 [4.12, 4.35] |
| 11 | 1.97 [1.89, 2.04] | 3.87 [3.74, 4.03] | 2.48 [1.98, 2.94] | 4.15 [3.93, 4.33] |
| 12 | 2.04 [1.97, 2.11] | 3.90 [3.75, 4.04] | 2.43 [1.91, 2.94] | 4.23 [4.08, 4.39] |
| 13 | 1.98 [1.92, 2.06] | 3.91 [3.77, 4.06] | 3.04 [2.48, 3.70] | 4.16 [4.03, 4.26] |
| 14 | 1.95 [1.89, 2.03] | 3.88 [3.73, 4.02] | 3.21 [2.69, 3.72] | 4.14 [3.99, 4.30] |
| 15 | 1.97 [1.90, 2.04] | 3.85 [3.72, 4.00] | 2.55 [1.94, 3.08] | 4.01 [3.86, 4.16] |
| 16 | 2.04 [1.97, 2.13] | 3.98 [3.83, 4.12] | 2.97 [2.25, 3.68] | 4.24 [4.12, 4.36] |
| 17 | 1.98 [1.91, 2.04] | 3.79 [3.65, 3.94] | 2.79 [2.21, 3.46] | 4.24 [3.99, 4.43] |
| 18 | 1.93 [1.87, 2.00] | 3.86 [3.74, 4.01] | 2.05 [1.18, 2.90] | 4.17 [4.04, 4.28] |
| 19 | 1.93 [1.87, 2.00] | 3.93 [3.77, 4.06] | 2.34 [1.75, 2.84] | 3.98 [3.83, 4.14] |
| 20 | 2.05 [1.97, 2.12] | 3.78 [3.65, 3.92] | 3.35 [2.84, 3.82] | 4.16 [4.06, 4.27] |

Table 3: Transmission rate estimates for all 20 simulation replicates of simulation settings V and VI (subsets of 10 non-identically distributed clusters) and their respective inference settings ('1 to 10': estimation of transmission rate of clusters 1 to 10, '11 to 20': estimation of transmission rate of clusters 11 to 20): Mean of the posterior sample and 95% highest posterior density interval in square brackets.

### SUPPLEMENTAL TABLE

**Data Availability**

| | |
|---|---|
| GISAID Identifier: | EPI_SET_250119mc |
| doi: | 10.55876/gis8.250119mc |

All genome sequences and associated metadata in this dataset are published in GISAID's EpiCoV database. To view the contributors of each individual sequence with details such as accession number, Virus name, Collection date, Originating Lab and Submitting Lab and the list of Authors, visit 10.55876/gis8.250119mc

**Data Snapshot**

- EPI_SET_250119mc is composed of 826 individual genome sequences.
- The collection dates range from 2020-01-28 to 2020-03-22;
- Data were collected in 1 countries and territories;
- All sequences in this dataset are compared relative to hCoV-19/Wuhan/WIV04/2019 (WIV04), the official reference sequence employed by GISAID (EPI_ISL_402124). Learn more at https://gisaid.org/WIV04.

Table 4: GISAID acknowledgements.
